# A longitudinal resource for studying connectome development and its psychiatric associations during childhood

**DOI:** 10.1101/2021.03.09.21253168

**Authors:** Russell H. Tobe, Anna MacKay-Brandt, Ryan Lim, Melissa Kramer, Melissa M. Breland, Lucia Tu, Yiwen Tian, Kristin Dietz Trautman, Caixia Hu, Raj Sangoi, Lindsay Alexander, Vilma Gabbay, F. Xavier Castellanos, Bennett L. Leventhal, R. Cameron Craddock, Stanley J. Colcombe, Alexandre R. Franco, Michael P. Milham

## Abstract

Most psychiatric disorders are chronic, associated with high levels of disability and distress, and present during pediatric development. Scientific innovation increasingly allows researchers to probe brain-behavior relationships in the developing human. As a result, ambitions to (1) establish normative pediatric brain development trajectories akin to growth curves, (2) characterize reliable metrics for distinguishing illness, and (3) develop clinically useful tools to assist in the diagnosis and management of mental health and learning disorders have gained significant momentum. To this end, the NKI-Rockland Sample initiative was created to probe lifespan development as a large-scale multimodal dataset. The NKI-Rockland Sample Longitudinal Discovery of Brain Development Trajectories substudy (N=369) is a 24- to 30-month multi-cohort longitudinal pediatric investigation (ages 6.0-17.0 at enrollment) carried out in a community-ascertained sample. Data include psychiatric diagnostic, medical, behavioral, and cognitive phenotyping, as well as multimodal brain imaging (resting fMRI, diffusion MRI, morphometric MRI, arterial spin labeling), genetics, and actigraphy. Herein, we present the rationale, design, and implementation of the Longitudinal Discovery of Brain Development Trajectories protocol.

## Background & Summary

Psychiatric disorders are common throughout the lifespan, imparting significant burden to individuals, families, and societies ^1–3^. At least 50% of psychiatric illnesses present before age 14 and 75% prior to age 24 ^4–7^. This has drawn attention to the first two decades of life for efforts to understand disruptions in brain development underlying the onset of illness and identify modifiable targets for intervention ^4–7^. Discrete periods of susceptibility for the emergence of psychiatric illness are known to exist within the developmental period, making longitudinal examinations spanning childhood and adolescence particularly valuable ^8–10^. For example, early childhood is characterized by the emergence of disruptive behavior, impulse control, and anxiety disorders, while adolescence is notable for the onset of mood, psychotic, and substance use disorders ^4, 11–15^.

Here, we provide an overview of a childhood-focused (enrollment ages 6-17) longitudinal, multimodal data resource created and openly shared by the NKI-Rockland Sample (NKI-RS) initiative—a resource explicitly created to accelerate the pace of scientific discovery for understanding brain development and the disruptions associated with the onset of psychiatric illness. While the NKI-RS core cross-sectional characterization brings numerous advantages, longitudinal investigations are required to delineate developmental trajectories, infer causality, and profile disorder incidence ^7^.

Key defining features in addition to the longitudinal nature include:

1) **Community-ascertained, transdiagnostic recruitment strategy.** The NKI-RS initiative is one of several neuroimaging efforts to amass large prospectively and openly shared datasets ^16^. Defining features of the NKI-RS are reliance on a community-ascertained design and broad inclusion criteria that allow for participants with mild-to moderate psychiatric illness (past or present), regardless of the specific nature of the psychiatric illness present. This strategy is highly consistent with the transdiagnostic agenda embodied in the NIH Research Domain Criteria Project and its counterparts ^17, 18^, and avoids the pitfalls of case-control recruitment strategies—which tend to restrict heterogeneity and yield “super-healthy” control samples ^17, 19–23^. Such heterogeneity is critical for characterizing complex brain-behavior relationships, many of which can be subtle, as well as establishing reference health measures and inferring causal connections in a longitudinal program ^19, 24^.
2) **Comprehensive psychiatric and behavioral phenotyping, including diagnostic assessments.** The NKI-RS maintains a shared core deep phenotypic battery to enable the association of multimodal imaging with extensive behavioral, physical, cognitive, psychological, and diagnostic characterizations. The depth of its phenotyping for mental health symptomatology, including substance use, differentiates its protocol from less psychiatrically focused contemporary studies, such as the NIH HCP-Development and ABCD studies ^25, 26^. By connecting brain structure and function to behavior, these rich neurobehavioral characterizations facilitate probing trajectories in the context of typical development, instances of behavioral symptoms, and occurrence of psychiatric diagnosis. While the cross-sectional lifespan nature of the aggregate NKI-RS and its large sample size position the dataset for developmental hypothesis generation, the longitudinal characterizations of a pediatric sub-cohort facilitate testing of those premises. Inclusion of widely used tools, such as the Child Behavior Checklist (CBCL) ^27^ and Kiddie Schedule for Affective Disorders and Schizophrenia (KSADS) ^28^, allows for the possibility of comparing, harmonizing, and aggregating results with other psychiatrically focused samples, such as the Philadelphia Neurodevelopmental Cohort and Brazilian High Risk Cohort ^29, 30^.
3) **Multimodal imaging (resting state fMRI, breath holding fMRI, diffusion MRI, morphometry MRI, arterial spin labeling).** Multimodal MRI-based imaging approaches allow us to map the brain’s structural and functional architecture ^31^. Inter- and intra-individual variation in this architecture holds promise in understanding health, development, and psychopathology when associated with other neurobehavioral phenotypic characterizations ^32, 33^. An explicit goal of the NKI-Rockland Sample imaging protocols was to combine multiple modalities in a single sample to enable multifaceted characterizations and perspectives of functional and structural architecture of the brain, with a particular focus on the connectome. In this regard, the primary protocols include diffusion imaging, resting state fMRI imaging, and morphometry MRI. Brief task fMRI scans were included to enable assessment of contrast-to-noise ratio (i.e., visual checkerboard) and neurovascular coupling (i.e., breath hold task).
4) **Multiband imaging.** The NKI-Rockland Sample was among the first initiatives to receive the multiband sequence (MB) after its preview in the Human Connectome Project ^34, 35^. Given the novel technology, two MB4-based functional sequences (resting state duration = 10 minutes) were included - one showcasing increases in temporal resolution and the other spatial (i.e., 3mm isotropic voxel size/TR=645ms; 2 mm isotropic voxel-size/TR=1400ms); a brief (2 minutes and 27 seconds) visual checkerboard stimulation task is included for each resolution to facilitate calculations of contrast to noise ratio. A traditional single-band EPI rest fMRI scan was included as well (resting state duration: 5 minutes), to enable comparisons with older acquisitions. Additionally, a breath hold task fMRI scan was included to facilitate assessments of age-related variation in neurovascular coupling (the higher spatial resolution MB4 scan was employed for this; breath hold duration = 4 minutes and 30 seconds). For diffusion imaging, a single MB4-based 137-direction scan was included as well.
5) **Inclusion of retest imaging sessions for participants.** Test-retest reliability is a critical prerequisite for longitudinal examination, as experimental error between longitudinal timepoints inherently limits our capacity to detect meaningful time-related change ^6^. While it is a longstanding practice for questionnaires and assessments (e.g., K-SADS) in psychiatry to be assessed for test-retest reliability, such examinations have gained a central focus in neuroimaging only in recent years ^36–40^. Depending on the specific modalities, data quantities, measures and brain system/areas examined, findings regarding reliability can vary, with the least known about advanced methods (e.g., multiband imaging). Compounding the challenge at hand, with rare exceptions ^41, 42^, these studies have been conducted in adults, leaving reliability in children largely unknown.
6) **Multicohort design.** A multicohort longitudinal approach was adopted given the expense and duration associated with implementing a single cohort design to sample our target enrollment age-range: 6-17 years ^7^. With a multicohort approach, developmental trajectories are more readily mapped while hypotheses of causality generated from correlative cross-sectional analyses can be confirmed in overlapping longitudinal cohorts. Our initial ambition was to achieve an ideal strict structured multicohort design, in which an equal number of individuals of each sex are recruited into each age-group, though as discussed in greater detail under *Sample Biasing and Representativeness* of our Methods, criteria were loosened somewhat over the course of the study to ensure recruitment feasibility.

The present Data Descriptor is intended to provide essential details regarding our project and protocol plans. We discuss lessons learned in recruitment, retention, data collection, and data distribution, which help to provide insights into the challenges that investigators face when trying to attain recruitment and design ideals (e.g., community-ascertained recruitment strategy, multi-cohort design) in imaging studies. We provide basic descriptions of the cohort including demographics, symptom heterogeneity, and attrition. We provide demonstrations of correlative analyses leveraging neurobehavioral, psychological, and developmental phenotyping. Finally, with respect to neuroimaging, we present motion features, structural and functional results as a function of demography, and profiles of differential scanning parameters.

## Methods

### Recruitment Strategy

The target enrollment for the Longitudinal Discovery of Brain Development Trajectories project was 384, with 369 recruited between December 2013 and November 2017. The last follow-up characterization visit was completed in June 2019. To best leverage statistical power and generalizability, a community-ascertained recruitment strategy was deployed to approximate a community-representative enrollment for the local region.

To accomplish recruitment and community engagement, the NKI-RS program staff targeted multiple educational and recruitment outreach opportunities in the region (Rockland County) to foster a community-collaborative approach. Primary referral pathways are summarized in Supplementary Table 1 for all participants. Successful partnerships with community members came predominantly from study recruitment booths at community events including local street fairs and the NKI Neuroscience Education Day (a.k.a. NKI Brain Day). The NKI Brain Day was developed in a collaborative educational effort with the NYU Neuroscience Department while leveraging and adapting materials from the Dana Foundation^43^. The overall goal was to provide a hands-on educational neuroscience experience for families. Activities included education about neuroimaging with a mock scan exposure, lectures, facility tours, a laboratory with models and animal brain specimens, optical illusions and other brain games, as well as a neuroscience art area. All components were led by study staff and provided parallel opportunities to familiarize potential participants with the program and facility. As the program grew, word of mouth became a prominent referral source. Regionally-targeted InfoUSA®-based mailers, email blasts, and appeals for word-of-mouth recommendations facilitated representation from areas that were relatively under-represented.

Given the scope and potential clinical applicability of the collected information, feedback was provided to all participant families by a licensed clinician. The feedback included pertinent physical measures, imaging findings and laboratory results (complete blood count, comprehensive metabolic panel, thyroid stimulating hormone, lipid profile, lead level) along with results from the semi-structured research diagnostic assessment and normed behavioral, cognitive, mood and anxiety measures. Notably, feedback was consistently presented as ‘screening’ with limited comprehensive clinical applicability in recruitment efforts. Mental health clinics, developmental pediatricians, neurologists, clinical psychologists, psychiatrists, and specialized school staff were not targeted as referral resources given concerns of generalizability of the sample; however, 5% of enrolled participant families learned of the study from a clinical provider (Supplementary Table 1).

### Retention Strategy

Several procedures and protocol adjustments were implemented to promote retention. These included:

- Birthday cards were sent to participants to help maintain a positive connection and track address changes.
- Enrolled participants were invited to educational talks and events designed to promote an understanding of the complexities of the brain and the need for brain imaging studies; preliminary findings and results were relayed as well. The scientific importance of the longitudinal dataset was emphasized to encourage retention.
- Written and verbal participant satisfaction was ascertained at all protocol visits to ensure the experience was regarded as positive and worthwhile at the participant and community level.
- Based on participant feedback, the baseline characterization protocol was compressed from two six-hour days to a brief consent visit easily arranged after school hours followed by a one day, 8-hour intensive protocol. This change took effect in mid-September 2015. Protocol changes are detailed in Supplementary Figure 1. In addition to decreasing the number of full site days, use of a brief consent visit limited the duration of participant fasting for phlebotomy because participants did not complete consent procedures while fasting.
- Retest MRI scans were permitted after any study visit rather than only the baseline characterization visit. Participants who failed to present for the midpoint visit were not withdrawn but asked to complete the final protocol visit. A maximal 3-month follow-up window was provided to facilitate scheduling of protocol visits.
- The initial protocol called for two longitudinal follow-up timepoints at 15 and 30 months following the baseline (referred to as: 0/15/30 track) to minimize possible season-related variation between participants. However, over the course of the study, the long duration between longitudinal visits became a source of concern regarding retention, leading to the creation of a complementary track with 12 months between consecutive visits rather than 15 (0/12/24 track). In addition to adding flexibility, the 0/12/24 month track helped families anchor annual longitudinal follow-up visits during a standard school break and shortened the study enrollment period.
- The availability of visits on non-school days was maximized in response to participant feedback; 87.6% of visits occurred on non-school days including 60.1% of visits on weekends, 23% during winter/spring/summer breaks, 4.1% on national holidays, and 0.4% on teacher conference days.

Of the 369 participants enrolled, 326 (88.3%) completed the baseline visits in their entirety. Among those with completed baseline visits, 247 (75.8%) completed the midpoint visit, 177 (54.3%) completed the final protocol visit, and 207 (63.5%) completed a retest visit (see Figure 1 for detailed breakdown).

**Figure 1.**
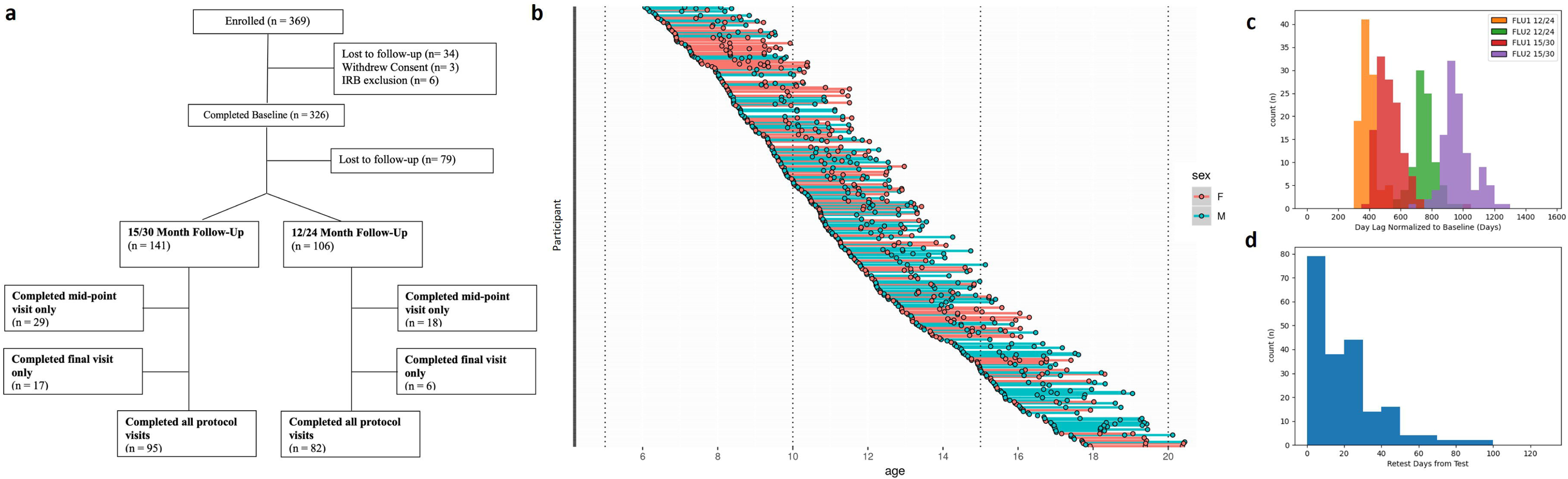
Participant enrollment and attrition. (a) Adapted CONSORT diagram demonstrating group level attrition by longitudinal follow-up track (either 0/12/24 month or 0/15/30 month follow-up) and characterization time-points, (b) Participant-level characterization time-point completion stratified by age at enrollment, females coded as red and males coded as blue, (c) Participant-level days to follow-up characterization visit from baseline, (d) Participant-level days to retest from most recent characterization visit. FLU1 = Follow-up Visit 1 (mid-point visit), FLU2 = Follow-up Visit 2 (final visit).

### Participant Procedures

#### Screening

To determine eligibility and ensure safety, legal guardians of potential participants completed a pre-screening phone interview with an intake coordinator. The screening interview inquired about the participant’s health history and assessed for study inclusion criteria (Table 1). Individuals meeting study criteria were invited to participate.

**Table 1.** Participant inclusion and exclusion criteria.

| Inclusion Criteria |
| --- |
| 1. Male or female ages 6.0-20.5 years (ages 6.0-17.9 at enrollment) |
| 2. Children who become adults during participation have capacity to understand and provide informed consent |
| 3. Children ages 6.0-17.9 with capacity to sign assent and parent/guardian with capacity to sign informed consent |
| 4. Fluent in English |
| 5. Proof of Rockland County, Orange County, Bergen County, or Westchester County Residency |
| Exclusion Criteria |
| 1. Parents unable to provide developmental and/or biological family histories (e.g., some instances of adoption) |
| 2. Known intrauterine exposures capable of altering brain structure or function (teratogenic medications, any illicit drug use, smoking $> \frac{1}{2}$ pack per day or $> 2$ alcoholic drinks per week during pregnancy), hyperbilirubinemia requiring transfusion or phototherapy $> 2$ days, multiple birth, infant resuscitation by chest compression or intubation, birth weight $< 1500$gm or $> 4200$ gm. |
| 3. Current height or weight $< 3$rd percentile, or head circumference $< 3$rd percentile by National Center for Health Statistics 2000 data. |
| 4. History of significant medical or neurological disorder with CNS implications (e.g., seizure disorder, CNS infection, malignancy, diabetes, systemic rheumatologic illness, muscular dystrophy, migraine or cluster headaches, sickle cell anemia, etc.), significant traumatic brain injury |
| 5. History of neonatal stroke |
| 6. Malignancy |
| 7. Hearing or visual impairment that prevents participation in study-related tasks. |
| 8. Current positive pregnancy test (for menarchal females) |
| 9. Current or past treatment for language disorders (except simple articulation disorders) |
| 10. History of clinically significant psychotic episode not attributable to a general medical condition or medication side effect |
| 11. History of Autism Spectrum Disorder |
| 12. Current suicidal/homicidal ideation |
| 13. History of treatment with the following psychotropic medications: antidepressants, neuroleptics, and mood stabilizers |
| 14. History of lifetime substance dependence requiring chemical replacement therapy |
| 15. History of substance dependence within in the last two years except nicotine and marijuana |
| 16. History of psychiatric hospitalization |
| 17. IQ < 70 |
| 18. Known neurodegenerative disorder |
| 19. First degree relatives with lifetime history of autism spectrum disorder, idiopathic intellectual disability, schizophrenia, psychotic disorder |
| 20. Contraindication for MRI scanning (metal implants, pacemakers, claustrophobia, metal foreign bodies or pregnancy) |

#### Medications

Participants with a history of antidepressant, mood-stabilizing, or neuroleptic medication use were excluded at enrollment (Table 1). Participants taking stimulant medications were asked to discontinue their medication during protocol visit days, as stimulant medications can affect cognitive and behavioral testing as well as functional brain mapping. Participants who chose not to discontinue medications, or whose physicians required that medication not be interrupted, were enrolled and completed all study procedures.

#### IRB approval

The study was approved by the Nathan Kline Institute for Psychiatric Research Institutional Review Board. Prior to conducting the research, written informed consent was obtained from participants’ legal guardians and written assent was obtained from the participants. Participants who became adults in the longitudinal follow-up provided written consent once becoming 18 years old.

### Experimental Design

Participants had 5 study visits. As highlighted in **Retention Strategy**, the baseline characterization included MRI and consisted of either two 6-hour visits if enrolled prior to mid-September 2015 or a brief consent visit followed by an 8-hour characterization visit if enrolled after. Mid-point and final visits were a single 8-hour characterization including MRI. The MRI retest visit was ideally within 3 weeks of either the baseline, mid-point, or final visit. All assessments are listed in Table 2. Participants were assigned to complete assessments in one of three characterization schedules. An example is provided in Table 3; see Supplementary Figure 2 and/or http://fcon_1000.projects.nitrc.org/indi/enhanced/CLGFullEndUserProtocol.pdf for full details.

**Table 2.** Complete protocol. † Can be completed as a home assessment. ‡ Optional at follow-up based on time availability.

| Assessments/Procedures | Baseline Visit | Follow-Up Visit |
| --- | --- | --- |
| **General** |  |  |
| Brain Injury Screening Questionnaire (BISQ)<sup>†</sup> | ✓ | ✓ |
| Child/Adult Satisfaction Questionnaire | ✓ | ✓ |
| Consent | ✓ | ✓ |
| Demographic Questionnaire | ✓ | ✓ |
| Demographics Supplement | ✓ |  |
| Family History Questionnaire – Parent | ✓ |  |
| Hollingshead Four-Factor Index of Socioeconomic Status (SES) | ✓ |  |
| Medical History Questionnaire – Child<sup>†</sup> | ✓ | ✓ |
| Medications/Medical Conditions – Parent<sup>†</sup> | ✓ | ✓ |
| **Physical** |  |  |
| 6-Minute Bike Test | ✓ | ✓ |
| Actigraphy | ✓ | ✓ |
| Blood Draw | ✓ | ✓ |
| Cambridge-Hopkins Restless Legs Syndrome (CHRLS, 18+)<sup>†</sup> |  | ✓ |
| Color Vision | ✓ |  |
| Edinburgh Handedness Questionnaire (EHQ, 6-17) <sup>†</sup> | ✓ |  |
| Grip Strength | ✓ |  |
| Grooved Pegboard | ✓ | ✓ |
| International Physical Activity Questionnaire (IPAQ, 15+)<sup>†</sup> | ✓ | ✓ |
| Pittsburgh Sleep Quality Index (PSQI, 13+)<sup>†</sup> | ✓ | ✓ |
| Tanner Scale | ✓ | ✓ |
| Weight/Height, Vital Signs, Hip/Waist Measurement | ✓ | ✓ |
| **Diagnostic** |  |  |
| Kiddie Schedule for Affective Disorders and Schizophrenia, Present and Lifetime Version (KSADS-PL, 6-17) | ✓ | ✓ |
| Structured Clinical Interview for DSM Disorders (SCID, 18+) |  | ✓ |
| Children's Yale-Brown Obsessive Compulsive Scale (CY-BOCS, 6-17) | ✓ | ✓ |
| Vineland Adaptive Behavior Scales (VABS, 6-17) | ✓ |  |
| Yale Global Tic Severity Scale (YGTSS, 6-17) | ✓ | ✓ |
| **Cognitive** |  |  |
| Attention Network Test (ANT) | ✓ | ✓ |
| Delis-Kaplan Executive Function System (D-KEFS) – Verbal Fluency (8+) | ✓ | ✓ |
| DKEFS – Trails (8+) | ✓ | ✓ |
| DKEFS – Design Fluency (8+) | ✓ | ✓ |
| DKEFS – Color-Word Interference Test (8+) | ✓ | ✓ |
| DKEFS – Tower (8+) | ✓ | ✓ |
| Digit Span | ✓ | ✓ |
| Penn Computerized Neurocognitive Battery (CNB) | ✓ | ✓ |
| Rey Auditory Verbal Learning Test (RAVLT; 8+) | ✓ | ✓ |
| Wechsler Abbreviated Scale of Intelligence-II (WASI) | ✓ |  |
| Wechsler Individual Achievement Test-II (WIAT) | ✓ |  |
| **Imaging** |  |  |
| Magnetic Resonance Imaging (MRI) Session | ✓ | ✓ |
| Mock Scan | ✓ |  |
| MRI-Questionnaire (MRIQ; 13+) | ✓ | ✓ |
| **Substance Use and Addiction Measures** |  |  |
| Urine Drug Test (11+) | ✓ | ✓ |
| Comprehensive Addiction Severity Index for Adolescents (CASI, 11+) | ✓ | ✓ |
| Fagerstrom Test for Nicotine Dependence (FTND, 18+) |  | ✓ |
| Modified Fagerstrom Tolerance Questionnaire – Adolescents (FTQA, 13-17) | ✓ | ✓ |
| National Institute on Drug Abuse Questionnaire (NIDA, 11+) | ✓ | ✓ |
| **Behavioral Tasks** |  |  |
| Behavioral Indicator of Resiliency to Distress (BIRD)<sup>‡</sup> | ✓ | ✓ |
| Dot Probe<sup>‡</sup> | ✓ | ✓ |
| **Behavior and Phenotypic Characterization Questionnaires** |  |  |
| The High-Functioning Autism Spectrum Screening Questionnaire (ASSQ, 6-17) † | ✓ | ✓ |
| Adult Self-Report (ASR, 18+) |  | ✓ |
| Adult Temperament Questionnaire (ATQ, 16+)† | ✓ | ✓ |
| Behavior Assessment System for Children 2<sup>nd</sup> Edition – Parent Rating Scale (BASC, 7-17) | ✓ | ✓ |
| Beck Depression Inventory (BDI, 18+) |  | ✓ |
| Child Behavior Checklist (CBCL, 7-17) | ✓ | ✓ |
| CBCL-Adaptive Functioning (7-17) | ✓ | ✓ |
| Children’s Behavior Questionnaire Very Short Form (CBQ-VSF, 6-8) † | ✓ | ✓ |
| Children’s Depression Inventory 2 (CDI-2, 11-17) | ✓ | ✓ |
| Child Eating Behavior Questionnaire (CEBQ, 7-11) † | ✓ | ✓ |
| Cognitive Failures Questionnaire (CFQ, 16+)† | ✓ | ✓ |
| Connors 3<sup>rd</sup> Edition – Parent Short Form (6-17) † | ✓ | ✓ |
| Connors 3<sup>rd</sup> Edition – Self-Report Short Form (8-17) † | ✓ | ✓ |
| Early Adolescent Temperament Questionnaire (EATQ, 9-15) † | ✓ | ✓ |
| Eating Disorder Examination Questionnaire (EDEQ, 13+)† | ✓ | ✓ |
| Inventory of Callous-Unemotional Traits – Parent Version (ICU-P, 7-17) † | ✓ | ✓ |
| Inventory of Callous-Unemotional Traits – Youth Self Report (ICU-Y, 13+)† | ✓ | ✓ |
| Interpersonal Reactivity Index (IRI, 13+)† | ✓ | ✓ |
| Multidimensional Anxiety Scale for Children (MASC, 8-17) † | ✓ | ✓ |
| NEO Five Factor Inventory - 3 (NEO, 12-17) † | ✓ |  |
| The 21-Item Peters et al. Delusions Inventory (PDI, 13+) | ✓ | ✓ |
| Phenotyping and eXposures (PhenX) Sex History Questionnaire (13+)† | ✓ | ✓ |
| Repetitive Behaviors Scale – Revised (RBS-R, 6-17) † | ✓ | ✓ |
| Sex Role Identity Scale (13+)† | ✓ | ✓ |
| Sexual Orientation Scale (13+)† | ✓ | ✓ |
| Social Responsiveness Scale-Parent Report (SRS, 7-17) † | ✓ | ✓ |
| State-Trait Anxiety Inventory (STAI, 18+)† |  | ✓ |
| Strengths and Weaknesses of Attention-Deficit/Hyperactivity Disorder Symptoms and Normal Behavior Scale (SWAN, 6-17) † | ✓ | ✓ |
| Three-Factor Eating Questionnaire (TFEQ, 12+)† | ✓ | ✓ |
| Trauma Symptom Checklist for Children (TSC-C, 11-17) | ✓ | ✓ |
| Trauma Symptoms Checklist for Adults (TSC-40, 18+) |  | ✓ |
| University of California at Los Angeles Posttraumatic Stress Disorder Reaction Index (UCLA-PTSD) Parent (6-17) | ✓ | ✓ |
| UCLA-PTSD Child (8-17) | ✓ | ✓ |
| UPPS-P Impulsive Behavior Scale (18+) |  | ✓ |
| Youth Self-Report (YSR, 11-17) | ✓ | ✓ |
| YSR Adaptive Functioning (11-17) | ✓ | ✓ |
| Youth Risk Behavior Surveillance System Middle School/High School (YSRBS, 12+) | ✓ | ✓ |

**Table 3.** Sample baseline characterization schedule. *During the cognitive testing blocks, breaks were provided between tasks at rater discretion as deemed necessary.

| Time (min) | Child Activity | Parent Activity |
| --- | --- | --- |
| 25 | Height, weight, vitals, hip/waist measurements, blood draw, genetics, Tanner staging, color vision, grip strength, medications and medical conditions collected, urine drug test (ages 11+), urine pregnancy test (postmenarchal females) | Tanner Staging (for children under 12) |
| 15 | Break/Breakfast | MRI Safety Screener (repeated) |
| 80 | MRI/MRIQ |  |
| 10 | Break |  |
| 60 | Penn Computerized Neurocognitive Battery* | KSADS (Parent Interview), Hollingshead SES |
| 20 | KSADS (Child Interview) | Vineland Behavior Scale |
| 30 | Break/Lunch |  |
| 120 | WASI, WIAT, DKEFS, RAVLT, Digit Span, Pegboard* |  |
| 15 | Break |  |
| 40 | Child onsite questionnaires | Parent onsite questionnaires |
| 30 | Attention Network Test |  |
| 15 |  | Family History questionnaire |
| 6 | 6-minute bike |  |
| 10 | Satisfaction Survey (Ages 12+) | Satisfaction Survey |

### Clinician-Administered Assessments

Clinician-administered assessments were obtained by a multidisciplinary team of psychologists, social workers, research nurses, and psychiatrists. All clinician-administered assessments were conducted by, or under supervision of, licensed clinicians. All cognitive assessments were administered by trained research staff under the supervision of a neuropsychologist. Participant responses were first scored by the administering rater with verification of scoring by another trained staff member. Separate psychiatric diagnostic and neuropsychology multidisciplinary team meetings occurred weekly to review administration, reconcile coding disparities, maintain consistent coding conventions, and minimize rater drift. Psychiatric diagnostic meetings generated consensus research diagnostic conclusions based on the results of the semi-structured research diagnostic summary combined with all other available information collected during the protocol (i.e., the best estimate method). Individuals for whom consensus and KSADS/SCID standard diagnostic summary were in agreement were coded as having no additional consensus diagnosis in the dataset to indicate the KSADS/SCID standard diagnostic summary was confirmed. In instances of discrepancy between consensus and KSADS/SCID standard diagnostic summary, the consensus diagnostic changes were specified.

#### Baseline Visit Only

*Wechsler Abbreviated Scale of Intelligence, Second Edition (WASI-II)* ^44^. All participants were administered the WASI-II during baseline assessment only. Four subtests: Vocabulary, Similarities, Block Design and Matrix Reasoning measure verbal comprehension and perceptual reasoning. The WASI-II subtest scores can be combined to estimate Full Scale Intelligence Quotient (FSIQ).

*Wechsler Individual Achievement Test (WIAT-IIA), Second Edition Abbreviated* ^45^. Three subtests were included: Word Reading ranging from phonological skills and letter recognition to word recognition; Numerical Operations ranging from counting and number recognition to complex calculations involving equations, fractions, decimals, etc.; and Spelling ranging from single and blended sound dictation to word dictation.

#### Repeating at Each Longitudinal Time Point

*Kiddie-Schedule for Affective Disorders and Schizophrenia (KSADS) for School-Aged Children Present and Lifetime Version (PL)* ^28^. All participants and their guardians were administered the KSADS-PL at baseline.

If they were under 18 throughout the study, they were administered KSADS-PL at mid-point and final visits. The KSADS-PL is a semi-structured DSM IV-based research diagnostic assessment.

*Structured Clinical Interview for DSM Disorders (SCID-I/NP)* ^46^. Participants who turned 18 while participating were administered the SCID at mid-point and/or final visits, if applicable, instead of the KSADS. The SCID is a semi-structured DSM IV-based research diagnostic assessment. Unlike the KSADS, the guardian was not an informant on the SCID.

*Delis-Kaplan Executive Function System (D-KEFS)* ^47^. The full DKEFS battery was administered to participants who were 8 years and older. The full battery was introduced June 2014 and shortened in January 2017 to include the following subtests: Verbal Fluency, Trails, Design Fluency, Color-Word Interference, and Tower, (see Supplementary Figure 1).

*Rey Auditory Verbal Learning Test (RAVLT)* ^48^. The RAVLT, a 15-item word list repeated over 5 learning trials and recalled after an immediate and twenty-minute delay, was administered to participants 8 years and older to characterize verbal learning and memory.

*Digit Span* ^49^. The WISC-R version of digit span was administered to all participants to characterize short term and working memory via verbal recitation of short strings of serially presented numbers, recalled either in the same order as presented aurally by the examiner or in backwards order.

### Magnetic Resonance Imaging (MRI)

All MRI scanning took place on the 3.0T Siemens TIM Trio located at the Nathan Kline Institute. A 32-channel head coil was used for all acquisitions in our sample. Details regarding the imaging protocols are presented in Table 4. In each imaging visit, participants were asked to complete two morphometric sequences (T1-weighted, T2-FLAIR), a Diffusion Tensor Imaging (DTI) sequence, six fMRI runs, and one arterial spin labeling (ASL) sequence.

**Table 4.**
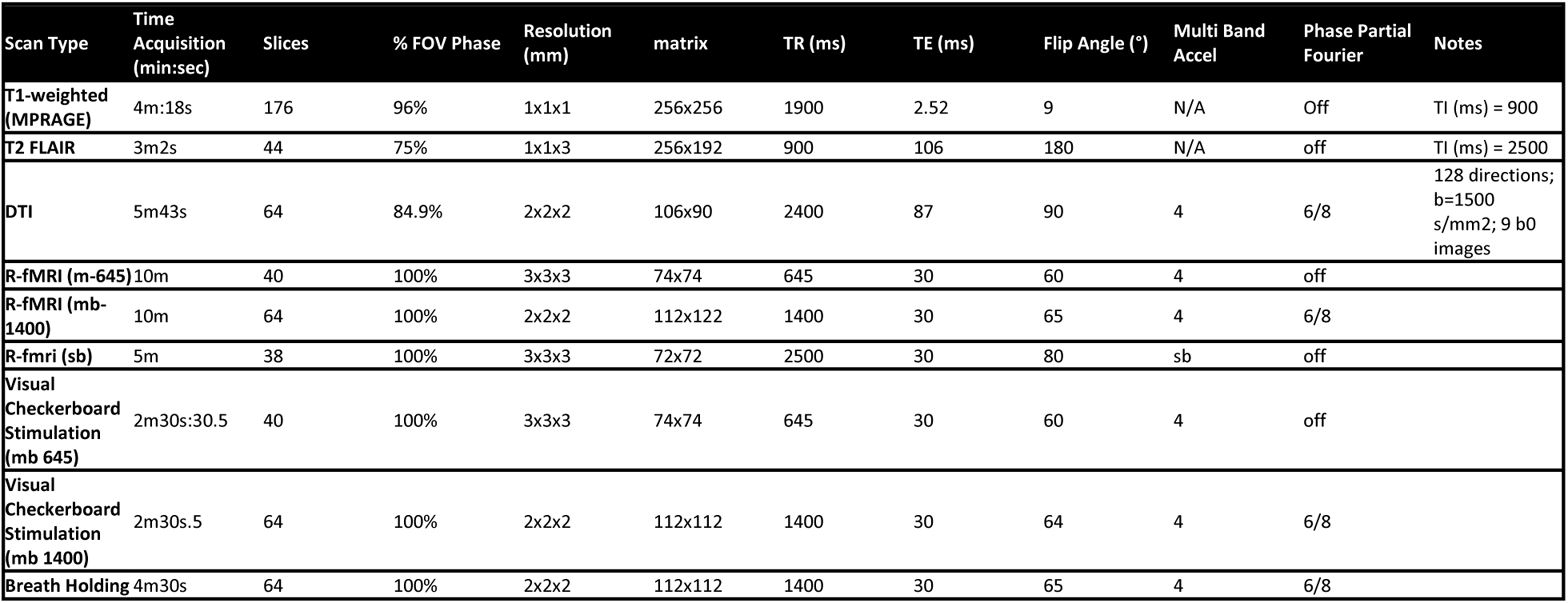
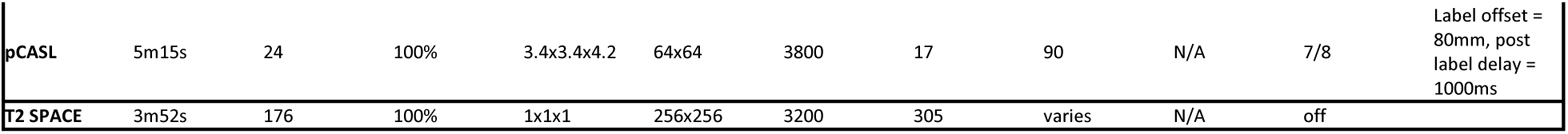
Imaging Protocol. * iso – Isotropic, * mb – Multi Band, * sb – Single Band, * pCASL – Pseudo-Continuous Arterial Spin Labeling. Full printouts of the imaging protocols can be downloaded here: http://fcon_1000.projects.nitrc.org/indi/enhanced/mri_protocol.html.

*Functional MRI.* For the six fMRI runs, three were resting state protocols, where the participants were asked to keep their eyes fixated on a crosshair, stay still, and not think of anything in particular. Two of these 10-minute resting state sequences used a multiband imaging protocol ^50^, one prioritizing temporal resolution (TR=645ms / 3mm isotropic voxels) and the other prioritizing spatial resolution (TR=1400ms / 2mm isotropic voxels). The third resting state sequence lasted 5 minutes and used typical single band acquisition parameters (TR=2500ms / 3mm isotropic voxels); it is included for reference. Two 2.5-minute visual checkerboard stimulation runs were completed by participants, one with each of the multiband sequences. The checkerboard task consisted of a flickering checkerboard (8Hz) in block design. Each of the 3 checkerboard blocks lasted 20 seconds, with 20-s resting periods between each stimulation block. A final breath-hold task was performed with a multiband sequence (TR=1400) for 4 minutes and 30 seconds. In this task, participants were given the following cues: “Rest,” “Get Ready,” “Breathe In,” “Breathe Out,” “Deep Breath and Hold.” Participants were then shown 6 circles of decreasing size, which indicated how much longer they needed to hold their breath. The “Rest” cue lasted 10 seconds; the “Get Ready,” “Breathe In,” “Breathe Out,” and “Deep Breath and Hold” cues each lasted 2.5 seconds; and the 6 circles each lasted 3 seconds (holding breath for 18 seconds in between). This cycle was repeated 7 times during the run.

*Diffusion Tensor Imaging.* DTI scans consisted of 128 diffusion directions with a b-value equal to 1500 s/mm2 and 9 b-0 images, with 2mm isotropic voxels, data collected in the AP direction, and with a multiband acceleration factor equal to 4. Acquisition time was 5 minutes and 43 seconds. During the scans participants listened to music.

*Morphometric Imaging.* Morphometric imaging consisted of T1-weighted and a T2-FLAIR scan. T1-weighted images were collected with 1mm3 isotropic voxels using an MPRAGE sequence. Each run took 4 minutes and 18 seconds to collect. A clinical T2-FLAIR was collected for radiological reading, with a slice thickness of 3mm and in plane resolution of 1mm^2^. Each run had a duration of 3 minutes and 2 seconds.

*Arterial Spin Labeling.* A pseudo-Continuous Arterial Spin Labeling (pCASL) sequence was run for each participant. As in the fMRI resting state sequences, participants were asked to keep their eyes fixated on a crosshair for 5 minutes and 15 seconds. The labeling plane offset was 80mm with a postlabeling delay (PLD) of 1s.

### Physical Assessment and Laboratory Analysis

Basic physical measurements (height, weight, and waist/hip circumference), cardiovascular measurements (heart rate and blood pressure), and grip strength were obtained by trained research staff under the supervision of team physicians and nurses at each of the three main protocol time points. Serum laboratory analysis was conducted at each main protocol time point: complete blood count, comprehensive metabolic profile, lipid profile, lead level, and thyroid function testing. Fasting was encouraged but changed to optional in July 2015, with fasting status coded for each phlebotomy encounter (Supplementary Figure 1). Urine Toxicology was obtained for participants aged 11 and older. Of the 326 baseline completers, 157 (48.2%) provided a blood sample. Of the 247 midpoint completers, 117 (47.4%) completed phlebotomy. Of the 177 final visit completers, 95 (53.7%) completed phlebotomy. Study cohort laboratory test results are displayed in Supplementary Figure 3, along with clinical laboratory-determined healthy ranges. A genetics sample was obtained once during any protocol visit and sent to the NIMH Genetics Repository for further processing, analysis, and sharing. Of the 326 baseline completers, 162 (49.7%) submitted a genetics sample. Finally, actigraphy was obtained for up to four weeks prior to the characterization visit using the Phillips Respironics Actiwatch 2 system (see Actigraphy section below).

### Data Distribution and Use

*Development of an End User Inquiry Mechanism.* As of 12/31/2020, 229 collaborative research sites have obtained access to the enhanced NKI-RS dataset. An inquiry response mechanism to ensure “End Users” appropriately understand and access data was developed and has been retained after study completion. Without an inquiry mechanism, the capacity to leverage the dataset may be impacted, directly threatening relevance and utilization. Accordingly, an End User Response Panel—composed of program investigators with expertise in phenotyping, clinical characterization, database management, and imaging—was developed. Since implementation, the Panel has received and jointly responded to 232 unique inquiry threads (2017: 83; 2018: 62; 2019: 41; 2020: 46) to.

### Summarizing Lessons Learned

Throughout program implementation, critical appraisal of strategic approaches in recruitment, retention, characterization, and dissemination to the scientific community was performed in an ongoing manner through weekly program leadership meetings. While large-scale research implementation is based on study-specific factors, some key considerations that may support similar efforts are listed below.

*Participant Engagement:* The NKI-Rockland Initiative initially relied on InfoUSA-guided mailings to the community as a primary recruitment tool (yield typically: 1-3% per 10,000 flyers mailed). However, such efforts were of limited value in recruiting individuals under 18.0 years old. Accordingly, promotion of community and scientific partnership became a core tenet of recruitment and retention in this longitudinal cohort. A multifaceted approach was employed as follows: 1) efforts were made to form relationships with community agencies in promoting the advancement of the regional area as a benchmark for longitudinal pediatric developmental neuroscience; 2) educational initiatives were brought to schools and anchored at NKI in the form of NKI Brain Day; and 3) active and past participants, who were invited to community outreach and educational events, often acted as impromptu ambassadors to other non-enrolled program attendees. Participant word of mouth became a primary referral pathway, particularly as the study progressed. Consistent with our goals of representativeness, the resultant sample was found to broadly represent Rockland County and its surrounding areas (see Supplementary Table 2).

*Balancing Experimental Needs Against Participant Burden:* The initial cross-sectional design of the NKI-RS initiative applied a two-day on-site characterization battery that facilitated active proctoring of all questionnaires in a group-psychometric testing area. Participant feedback following direct translation of the two-day NKI-RS characterization to the longitudinal cohort suggested a need for consolidation to a one-day intensive characterization. Families cited the preference for weekend visits and minimal disruption to schooling as the study was not of clinical benefit to most participants. Accordingly, a brief consent visit was arranged and typically scheduled after school hours. Families were also given the option of completing questionnaires that did not assess clinically-defined safety items at home via the COllaborative Informatics and Neuroimaging Suite (COINS) web-based entry platform ^51, 52^. Participants and families were coached on minimizing distraction during home-based questionnaire entry. The remaining procedures from the initial two onsite 6-hour study visits were then consolidated to a single 8-hour visit. With this change, 87.6% of characterization visits were completed on non-school days with ample breaks and staggering of procedures to decrease data loss.

*Selection of Phenotypic Measures for Population Samples.* A substantial portion of the dimensional mental health assessments in the NKI-Rockland phenotypic battery is focused on detecting and quantifying psychiatric symptomatology (e.g., Conners ADHD Scale, Children’s Depression Inventory). Although reflective of the practices of the larger field, the reliance on measures that assess only the presence of deficits can be limiting as they fail to differentiate behaviors among unaffected individuals. Recent efforts by our team ^53^ and others ^54–57^ have called this practice into question as it is akin to grouping all with an IQ above 100 together. The distributions resulting from such tools tend to be truncated and thus suboptimal for dimensional analysis in genetics, imaging, and epidemiologic studies. The inclusion of both the Conners and SWAN for the assessment of ADHD—the former focused on detection of symptoms and the latter on that of strengths as well as weaknesses—allows interested users to explore this issue, which is not unique to the NKI-RS phenotypic battery. When possible, attempts were made to phenotype shared developmental constructs that extend across the larger population, e.g., Child Behavior Checklist ^27^, Early Adolescent Temperament Questionnaire ^58^, Adult Temperament Questionnaire ^59^, Penn Computerized Neurocognitive Battery ^60^, Vineland Adaptive Behavior Scales ^61^, and NEO Five Factor Inventory ^62^. However, given a lack of assessments emphasizing strength in psychiatric symptom characterization, standard clinical measures remained a central aspect of phenotyping. As the psychometric instruments producing dimensional distributions in non-clinical samples continue to become available, we encourage use of such measures in phenotyping investigations of community samples.

*Actigraphy.* Collection of actigraphy data was complicated by rates of unit damage and loss; 5 out of 28 units remained viable at the end of the project period. From a practical perspective, the number of units included in the budget at the onset of the project substantially underestimated real needs once the number of participants and visits per participant involved in the design were considered. Delays in visits at times caused stresses on our system due to the limited number of actigraphy units. The result of these challenges is that actigraphy represents one of the most notable sources of missing data in our sample. Future efforts would benefit from more careful estimation of the number of devices required to support the various contingencies that can arise in such a design. Recent actigraph price reductions increase feasibility of obtaining actigraphy data in large community studies. Participant incentives should be considered for consistent actigraph usage and return of functioning devices.

*Multicohort Design.* As highlighted previously, an initial structured multicohort design transitioned to an unstructured approach due to greater challenges of recruiting adolescents. While maintaining many strengths of a multicohort design, an unstructured design trades further flexibility in opportunistic recruitment for sample non-uniformity that may decrease (1) precision in delineation of age and cohort effects and (2) accuracy of mapping developmental trajectories ^7^. Pubertal-age participants (enrollment age 11 and up) and their families were more likely to cite conflicting demands (e.g., school activities, schoolwork, peer social networks, and summer camps) that often required high levels of commitment from the participant. Younger participants were more likely to complete all protocol visits (Age less than 11 = 60%, Age 11 and up = 48%, p = 0.03). Together, these observations may support the possibility that decreased assessment burden spread over more visits of shorter duration may facilitate improved adolescent recruitment and retention.

*Retention.* As discussed in the **Retention Strategy** section, several changes were made to provide protocol flexibility: (1) addition of a 12/24-month follow-up pathway; (2) consolidation of the baseline 2-day on-site characterization to a one-day intensive visit; (3) obtaining the retest MRI scans after any of the three time point visits; (4) providing a 3-month window for return visits; and (5) retaining participants who missed the midpoint visit. While these adaptations facilitated a larger and more complete dataset, they have several implications. (1) The 12/24 month follow-up pathway may be prone to seasonal developmental effects (e.g., probing a participant’s psychiatric symptoms only during their summer break); this may be a relevant consideration when pooling data from the 15/30-month follow-up pathway or directly considering seasonal effects. (2) Though other work by our group ^53^ has supported use of more frequent, shorter visits in clinical populations, this option was not provided given the larger non-clinical population. This approach also avoided confounds with variable characterization protocols. Participants who completed home-based assessments may have experienced conflicting attentional demands. (3) The ability to complete the retest at a later longitudinal visit inherently shifts the age distribution while likely improving data quality, with older participants suffering less motion artifact. (4) The 3-month visit window augments time effects in the sample as either a confounding variable or one for scientific exploration. (5) Retained participants with lesser protocol adherence may differ in background, family strain, or symptom measures not fully characterized by the protocol.

## Data Records

### Dataset deposition

Data files are accessible via the International Neuroimaging Data-Sharing Initiative (INDI) under the title “Enhanced Nathan Kline Institute - Rockland Sample (NKI-RS)” ^63^.

### Data privacy

During the consent process, all participants provide informed consent for their data to be shared via IRB-approved protocols. Given the sensitive nature of the information provided during NKI-RS participation, a Certificate of Confidentiality was obtained from the Department of Health and Human Services (HHS). The certificate helps to protect the privacy of human subjects by allowing the research team to refuse to disclose names or other identifying characteristics of study participants in response to legal demands (https://humansubjects.nih.gov/coc/index).

With respect to data-sharing via the International Neuroimaging Data-sharing Initiative (INDI), informed consent for sharing was obtained prior to participation in the study. Given the critical priority of protection of participant privacy, no protected health information is ever released via the INDI. For imaging data, face information was removed from anatomical scans, and protected/identifying information was removed from all image headers and databases. Additionally, random 7-digit INDI identifiers were assigned to all datasets. Recognizing the risks associated with sharing phenotypic data, which is high dimensional and personal in nature, we developed two release versions:

#### NKI-Rockland Lite

This release version contains only de-identified imaging data and limited phenotyping (age, sex, and handedness). No psychiatric, cognitive, or behavioral information is included. The NKI-Rockland Lite is immediately available without data usage agreement requirements. To access the NKI-RS Lite release, please visit the Study Website.

#### NKI-Rockland Full Phenotypic Release

This release contains the full high-dimensional phenotypic protocol. Phenotypic data includes comprehensive psychiatric and behavioral assessment questionnaires (item-level information included), neuropsychological testing, cognitive/behavioral performance measures, basic laboratory measures, respiratory and cardiac recordings obtained during imaging, and actigraphy (for a minimum 1 night of sleep). Users of the NKI-Rockland Full Phenotypic Release complete the following agreements and obtain appropriate institutional approval prior to accessing the data (expected completion time < 5 minutes). The entire phenotypic dataset will be made available to approved users via the COINS databasing system, which was the primary support for the NKI-Rockland data collection; also Longitudinal Online Research and Imaging System (LORIS) database, in which a curated version of the data has been created ^64^.

### Data Acknowledgment

A guiding principle for data-sharing in the NKI-Rockland Sample initiative is to provide researchers with the greatest flexibility to carry out scientific inquiries. In this regard:

1. We do not require authorship on any manuscripts generated using NKI-Rockland datasets; citation as a data-source is sufficient.
2. We do not require review of manuscripts generated using NKI-Rockland datasets; we do strongly encourage careful documentation of selection criteria applied to the NKI-RS in choosing datasets for analysis.
3. We do not require specification of data analyses prior to data usage.

### Distribution for use

The primary website for all NKI-Rockland Initiatives, including the present, is located at: http://fcon_1000.projects.nitrc.org/indi/enhanced/index.html. Phenotypic data may be accessed through either an NKI-RS-dedicated instance of the Longitudinal Online Research and Imaging System (LORIS) located at https://data.rocklandsample.rfmh.org/ and supported by the NKI-RS research program staff or through the COINS Data Exchange (https://coins.trendscenter.org/). Except for age, sex, and handedness, which are publicly available with the imaging, NKI-RS phenotypic data are protected by a Data Usage Agreement (DUA). Investigators must complete the DUA (found at http://fcon_1000.projects.nitrc.org/indi/enhanced/data/DUA.pdf) and have it approved by an authorized institutional official before receiving access. The intent of the DUA is to ensure that data users (1) agree to protect participant confidentiality when handling the high dimensional NKI-RS phenotypic data (which includes single item responses) and (2) agree to take the necessary measures to prevent breaches of privacy. The DUA does not place any constraints on the range of analyses that can be carried out using shared data, nor does it include requirements for co-authorship by the originators of the NKI-RS.

### Imaging data

All imaging data can be accessed through the 1,000 Functional Connectomes Project and its International Neuroimaging Data-sharing Initiative (FCP/INDI) based at http://fcon_1000.projects.nitrc.org/indi/enhanced/neurodata.html. This website provides an easy-to-use interface with point-and-click download of datasets that have been previously compressed; it also provides directions for users who are interested in direct download of the data from an Amazon Simple Storage Service (S3) bucket. A CSV file with the complete list of S3 links is provided in the study website. However, if a more specific list of S3 links is required (e.g., sex=female & age<10), it can be generated though LORIS. Imaging data is stored in the Brain Imaging Data Structure (BIDS) format, which is an increasingly popular approach to describing MRI data in a standard format. All data are labeled with the participant’s unique identifier.

### Partial and missing data

Some participants may not be able to successfully complete all components of the NKI-RS protocol due to a variety of factors (e.g., participants experiencing claustrophobia may not be able to stay in the scanner for the full session). To prevent data loss, when possible, we included a mock MRI scanner experience during the first visit. Overall, we attempt to collect as much of the data as possible within the allotted data collection intervals and log data losses when they occur.

### Data license

NKI-Rockland Sample data are distributed using the Creative Commons-Attribution-Noncommercial license, which is described at: https://creativecommons.org/licenses/by-nc/4.0/legalcode. For the high-dimensional phenotypic data, all terms specified by the DUA must be complied with.

## Technical Validation

### Sample Composition

#### Geographic Distribution

The Nathan S. Kline Institute for Psychiatric Research (NKI) is in Rockland County, NY, approximately 25 miles north of Manhattan. Outside of New York City, Rockland County is geographically the smallest New York county while housing the 8^th^ largest population at 311,687 ^65^. Residents of neighboring counties (Bergen County, NJ; Orange County, NY; and Westchester County, NY) were enrolled to facilitate total project enrollment goals and broaden demographic heterogeneity. However, Rockland County comprised most enrollments at 84%. Rockland County census data (Table 5) mirror that of the United States in most domains. Efforts were made to obtain proportionate representation for most zip code regions in Rockland County (Supplementary Table 2), however, communities in closer geographic proximity to NKI were more likely to enroll. A zip code region within Rockland county (10952, 10977) was notably underrepresented. Many residents in this region are members of a religious community that is found at a much higher rate in Rockland County than in the US population (Supplementary Table 2).

**Table 5.**
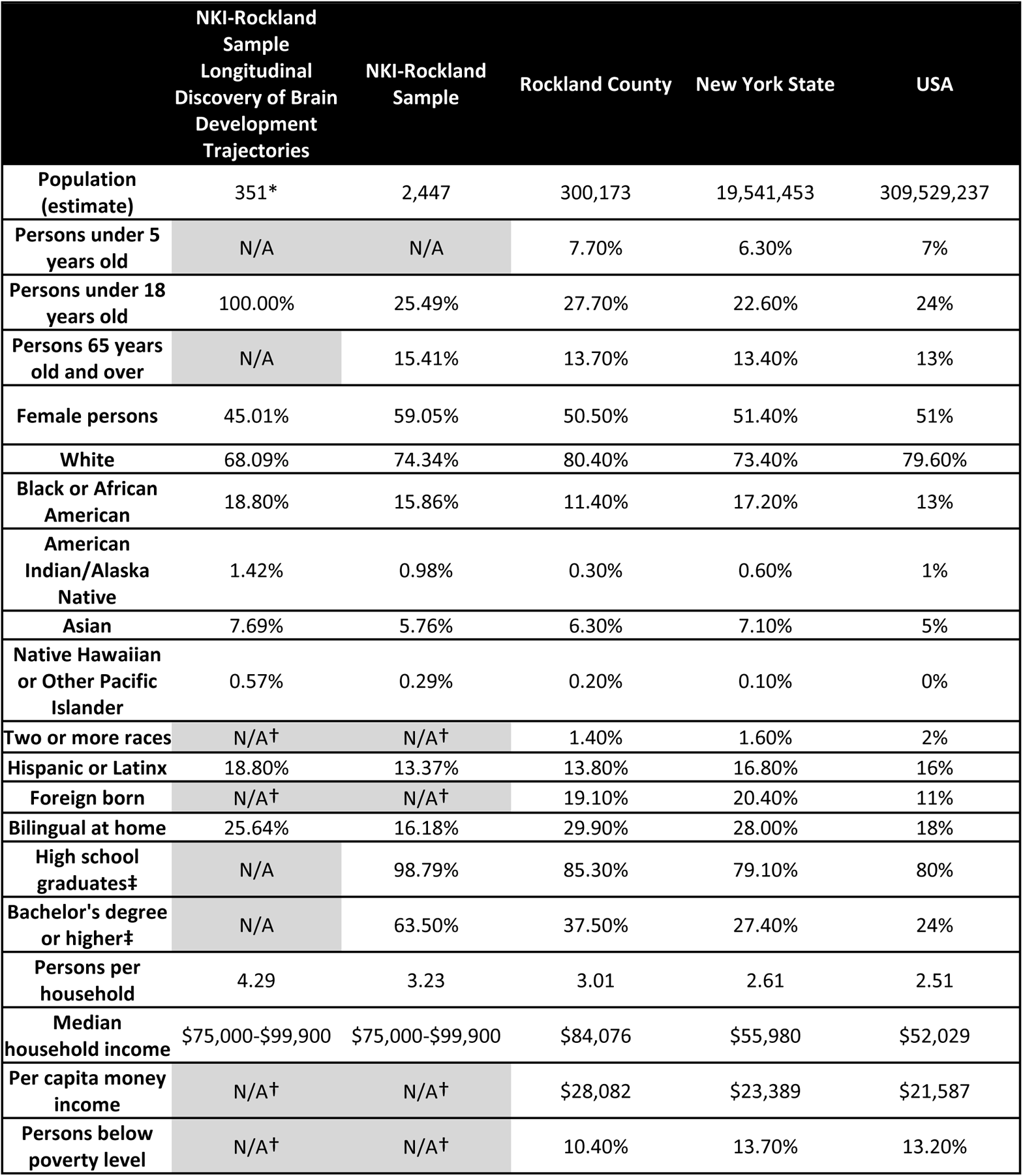
Demography of the NKI-Rockland Sample Longitudinal Discovery of Brain Development Trajectories sub-study (enrollment years 2013-2017); the NKI-Rockland Sample (enrollment years 2011-2019); along with the 2009 Rockland County, New York State, and United States Census Data. *Of the 369 participants enrolled, 351 participants completed baseline demography. †Several census variables were not replicated in the NKI-Rockland Sample: there was no participant option to identify two or more races or birth outside of the United States, and income was obtained as a range which could not be translated to per capita income or poverty thresholds. ‡Participants aged 25 and older.

|  | NKI-Rockland<br>Sample<br>Longitudinal<br>Discovery of Brain<br>Development<br>Trajectories | NKI-Rockland<br>Sample | Rockland County | New York State | USA |
| --- | --- | --- | --- | --- | --- |
| **Population (estimate)** | 351* | 2,447 | 300,173 | 19,541,453 | 309,529,237 |
| **Persons under 5 years old** | N/A | N/A | 7.70% | 6.30% | 7% |
| **Persons under 18 years old** | 100.00% | 25.49% | 27.70% | 22.60% | 24% |
| **Persons 65 years old and over** | N/A | 15.41% | 13.70% | 13.40% | 13% |
| **Female persons** | 45.01% | 59.05% | 50.50% | 51.40% | 51% |
| **White** | 68.09% | 74.34% | 80.40% | 73.40% | 79.60% |
| **Black or African American** | 18.80% | 15.86% | 11.40% | 17.20% | 13% |
| **American Indian/Alaska Native** | 1.42% | 0.98% | 0.30% | 0.60% | 1% |
| **Asian** | 7.69% | 5.76% | 6.30% | 7.10% | 5% |
| **Native Hawaiian or Other Pacific Islander** | 0.57% | 0.29% | 0.20% | 0.10% | 0% |
| **Two or more races** | N/A† | N/A† | 1.40% | 1.60% | 2% |
| **Hispanic or Latinx** | 18.80% | 13.37% | 13.80% | 16.80% | 16% |
| **Foreign born** | N/A† | N/A† | 19.10% | 20.40% | 11% |
| **Bilingual at home** | 25.64% | 16.18% | 29.90% | 28.00% | 18% |
| **High school graduates‡** | N/A | 98.79% | 85.30% | 79.10% | 80% |
| **Bachelor's degree or higher‡** | N/A | 63.50% | 37.50% | 27.40% | 24% |
| **Persons per household** | 4.29 | 3.23 | 3.01 | 2.61 | 2.51 |
| **Median household income** | \$75,000-\$99,900 | \$75,000-\$99,900 | \$84,076 | \$55,980 | \$52,029 |
| **Per capita money income** | N/A† | N/A† | \$28,082 | \$23,389 | \$21,587 |
| **Persons below poverty level** | N/A† | N/A† | 10.40% | 13.70% | 13.20% |

#### Diagnosis, Age and Sex

With respect to diagnosis in the 326 baseline completers, though fewer females enrolled (Figure 2a), numbers with no diagnosis (n=160) were comparable between the sexes (male = 78, female = 82, p = 0.752) indicating that boys in the sample were more likely to have at least one diagnosis (male = 98, female = 56, p < 0.001) (Figure 2b). Much of this difference was driven by high rates (25%) of ADHD diagnosis at baseline (male = 54, female = 25), which were, as expected, more frequent in boys (p < 0.001).

**Figure 2.**
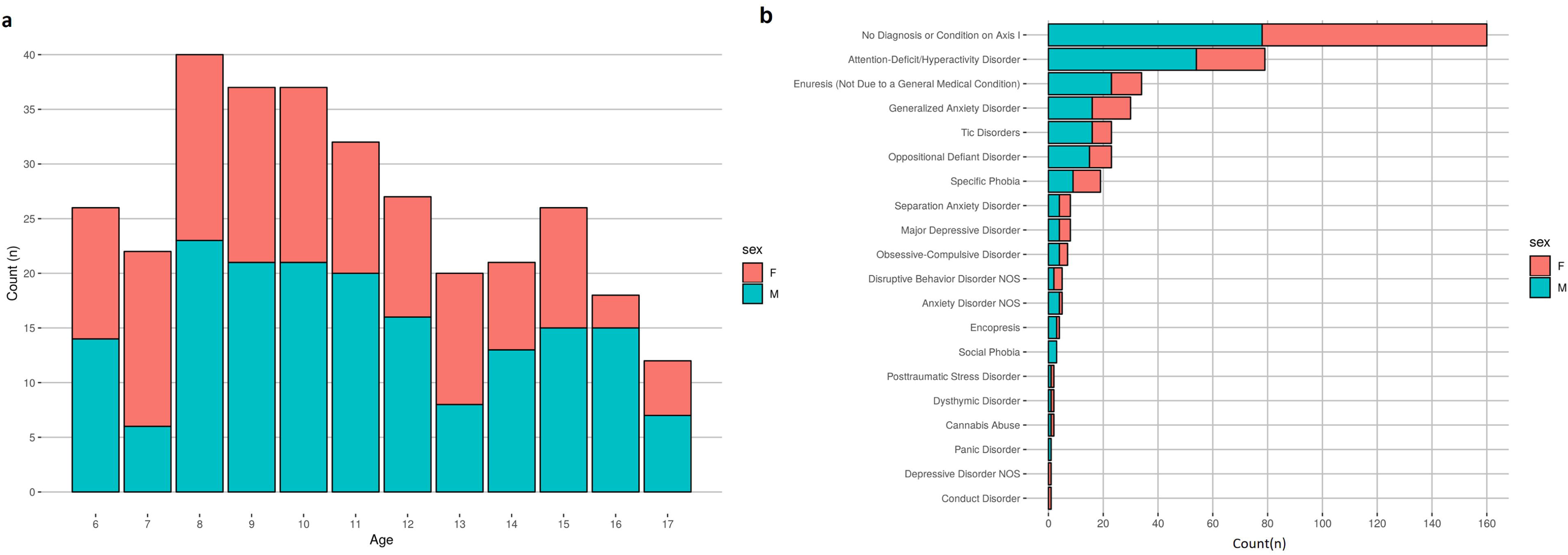
Age, sex, and diagnostic distribution of participants. (a) Age and sex distribution of participants. (b) Frequency of diagnoses given to participants during the baseline characterization visit. Participants could receive more than one diagnosis. Diagnostic data for this figure represent the ‘consensus diagnosis’ factoring all data collected during the protocol visit, including the K-SADS, medical history, and clinically-relevant measures. Females are coded as red, and males are coded as blue.

Rates of pediatric ADHD in our sample were above other epidemiologic samples of 9.4% ^66^. However, rates of other diagnoses were generally consistent with established community prevalence studies ^67^. Study-specific criteria excluded participants with history of psychiatric hospitalization, current suicidal/homicidal ideation as well as antidepressant, neuroleptic, or mood stabilizer use. As a result, psychiatric symptom severity at clinical thresholds warranting treatment was generally identified and excluded. However, use of ADHD medication was not exclusionary allowing enrollment of participants with clinically significant ADHD symptoms and families seeking refined assessment and guidance. We believe this was a major factor in selective elevated rates of ADHD in the sample.

#### Substance and Medication Use

Among the 326 baseline completers, 160 were aged 11 and up at baseline, with 158/160 completing substance use measures. 25, 9, 18, and 1 reported past alcohol, tobacco, marijuana, and other illicit substance use, respectively. All 160 submitted urine toxicology samples; 8 (7 for THC and 1 for amphetamine) had a positive urine toxicology for an illicit substance not readily explained by medications (i.e., stimulant medication in context of amphetamine positive urine). Rates of substance-related diagnosis (which was limited to cannabis abuse) are represented in Figure 2b. With respect to psychotropic medication use, many medication classes were excluded (see Table 1). 37 of the 351 guardians reported their child took any medication, 15 for behavioral health indications and 25 for medical indications which was most commonly asthma (n = 11). The majority of participants who took behavioral health medications did so in treatment of ADHD (n = 12; 11 took stimulants and 2 used non-stimulants).

#### Quality assessment

Consistent with policies established through our prior data generation and sharing initiatives, all imaging datasets collected through the NKI-RS are made available to users regardless of data quality given a lack of consensus on what constitutes “good” or “poor” quality data. Also, “lower quality” datasets can facilitate the development of artifact correction techniques and help in evaluating the impact of such real-world confounds on reliability and reproducibility.

#### Phenotypic data

Raw data were evaluated for technical errors prior to each release by reviewing scatterplots by age and boxplots comparing past releases to each progressive release. Additionally, group level data were inspected to ensure the observed distributions and inter-relationships were sensible. In keeping with the rationale for imaging data, phenotypic data are not ‘cleaned’ of outliers, unless data are identified in cursory reviews as containing overt technical errors. Data cleaning and review is expected to be incorporated into each independent research analysis. Figures 3–6 highlight expected performance on several core characterization measures. Figures 3 and 4 depict examples of assessments with symmetric (near normal) distributions (e.g., IQ, academic achievement testing, and SWAN) and truncated (e.g., CBCL, Conners) distributions. Shapiro-Wilk normality tests displayed in Figure 4 highlight relative differences between symptom-oriented (CBCL and Conners) attentional measures and the more normally distributed SWAN that emphasizes attentional strength in addition to symptom-associated weakness (see *Selection of Phenotypic Measures for Population Samples* section). This is further demonstrated by stronger subscale correlation trends between SWAN and Conners for participants with SWAN >= 0 (indicating more weakness as a proxy for symptomatology) over those with SWAN < 0. Figure 5 demonstrates anticipated improvement in both raw speed and errors with age for the Color Naming, Inhibition, and Switching conditions of the D-KEFS Color Word Interference Test. As expected, based on prior D-KEFS norming studies, standardized speed and error scores do not demonstrate prominent age effects aside from relatively under-recruited age bands. Figure 6 shows expected age-relationships in physical maturation including age-related improvements in motor speed/coordination (grooved pegboard) and strength (grip strength). Muscle bulking during pubertal onset as measured by Tanner stage coincides with increasing strength for boys relative to girls (Figure 6D) without differences in speed/coordination (Figure 6c) ^68–71^.

**Figure 3.**
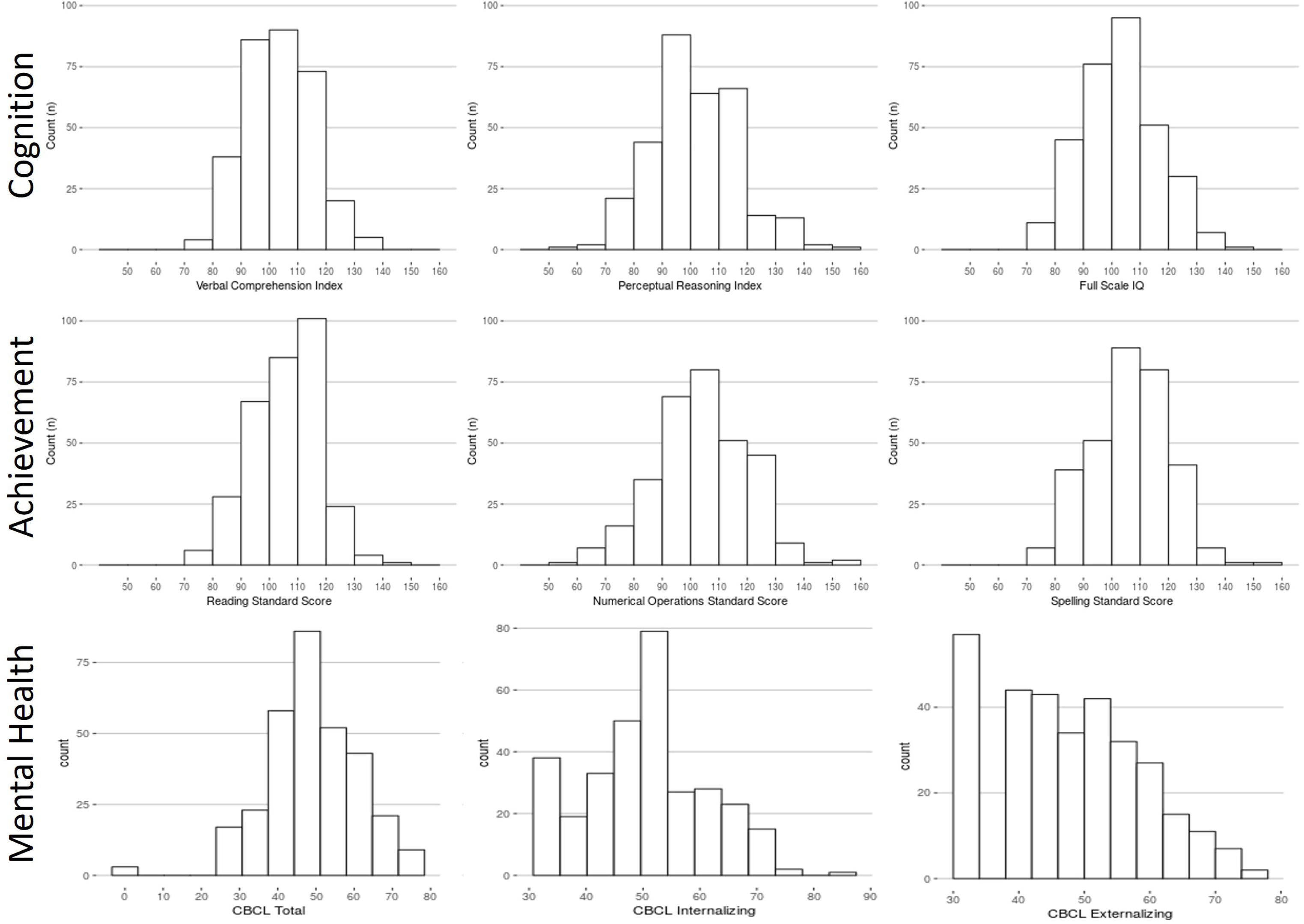
Distribution of general intellectual ability, academic achievement, and broad mental health measures. Participant IQ was estimated using the WASI-II (Top row left to right; (a) Verbal Comprehension Index Composite, (b) Perceptual Reasoning Index Composite, and (c) Full-Scale IQ Composite). Academic Achievement as measured by WIAT-II (Middle row left to right; (d) Word Reading, (e) Numerical Operations, and (f) Spelling Standard Scores). Mental Health as measured by CBCL (Bottom row left to right; (g) CBCL Total Score, (h) Internalizing Problems Scale, (i) Externalizing Problems Scale). Data are represented for all participants at baseline.

**Figure 4.**
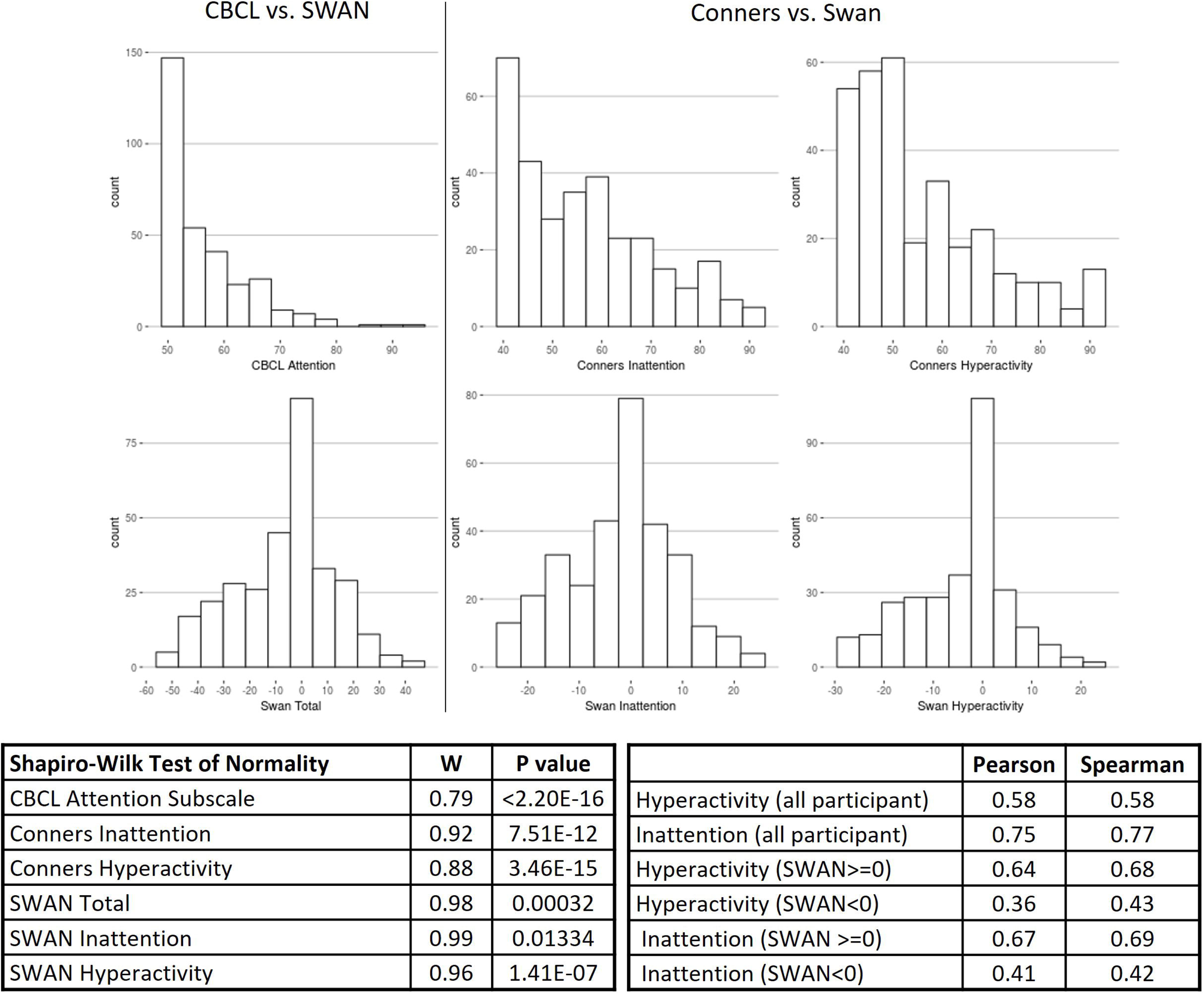
Distribution of attention ratings. Total CBCL Attention subdomain distribution compared to total SWAN distribution (left column). Distributions of Conners Inattention compared to SWAN Inattention (middle column) and Conners Hyperactivity compared to SWAN Hyperactivity (right column). The distribution of each scale was tested for normality using the Shapiro-Wilk test. Finally, Pearson (r) and Spearman (p) correlations between the SWAN and Conners are represented for both the hyperactivity and inattention subscales for all participants, those with SWAN >= 0, and participants with SWAN <0.

**Figure 5.**
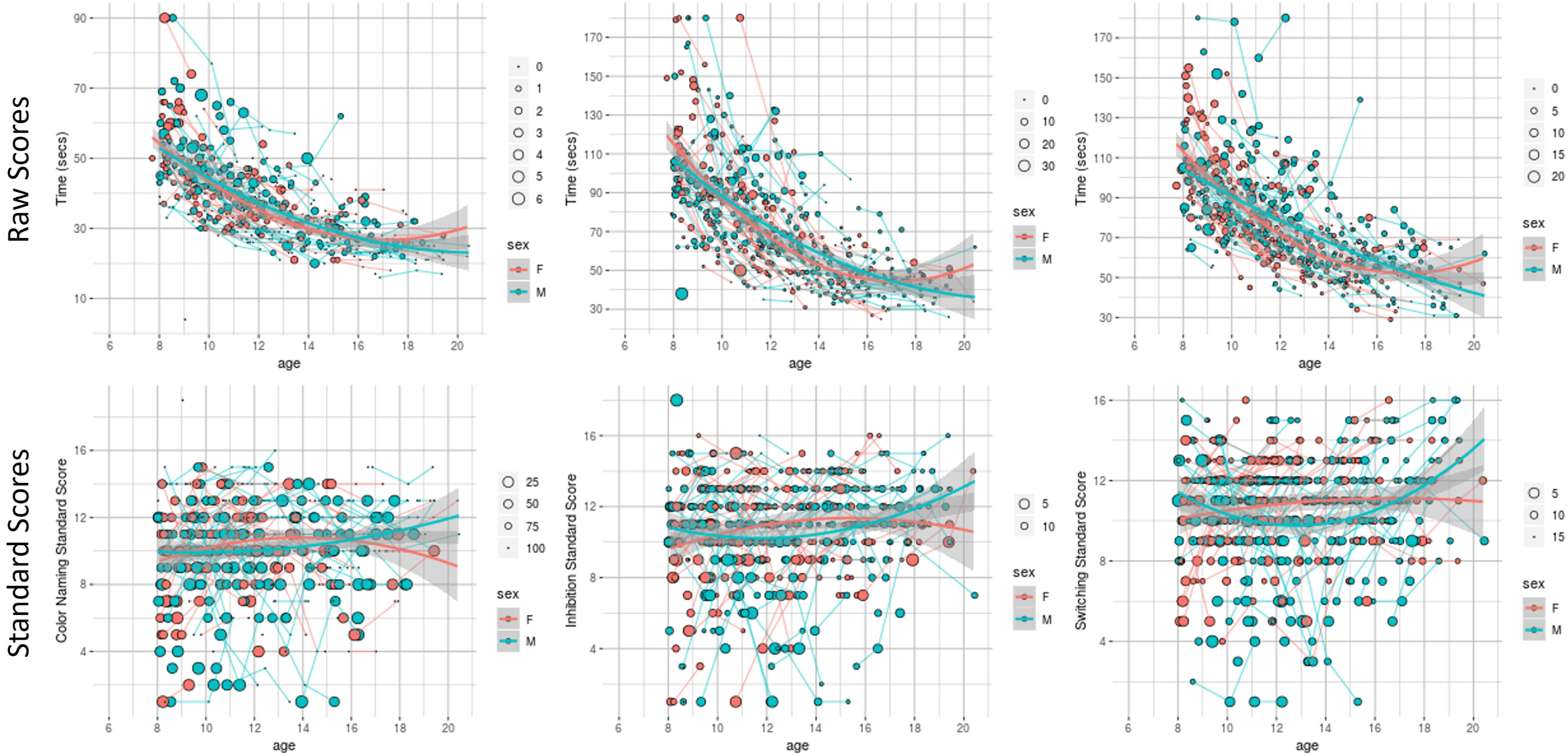
Raw and standardized results of the D-KEFS Color-Word Interference Test. (a,d) D-KEFS Color Word Interference Test Color Naming, (b,e) Inhibition, and (c,f) Switching conditions (from left to right) are represented by age for participants (ages 8+) with females coded as red and males coded as blue. Individual lines demonstrate participant-level longitudinal change. Raw scores and error are reflected in Row 1 while standard scores and error are reflected in Row 2. Larger circle sizes indicate more errors.

**Figure 6.**
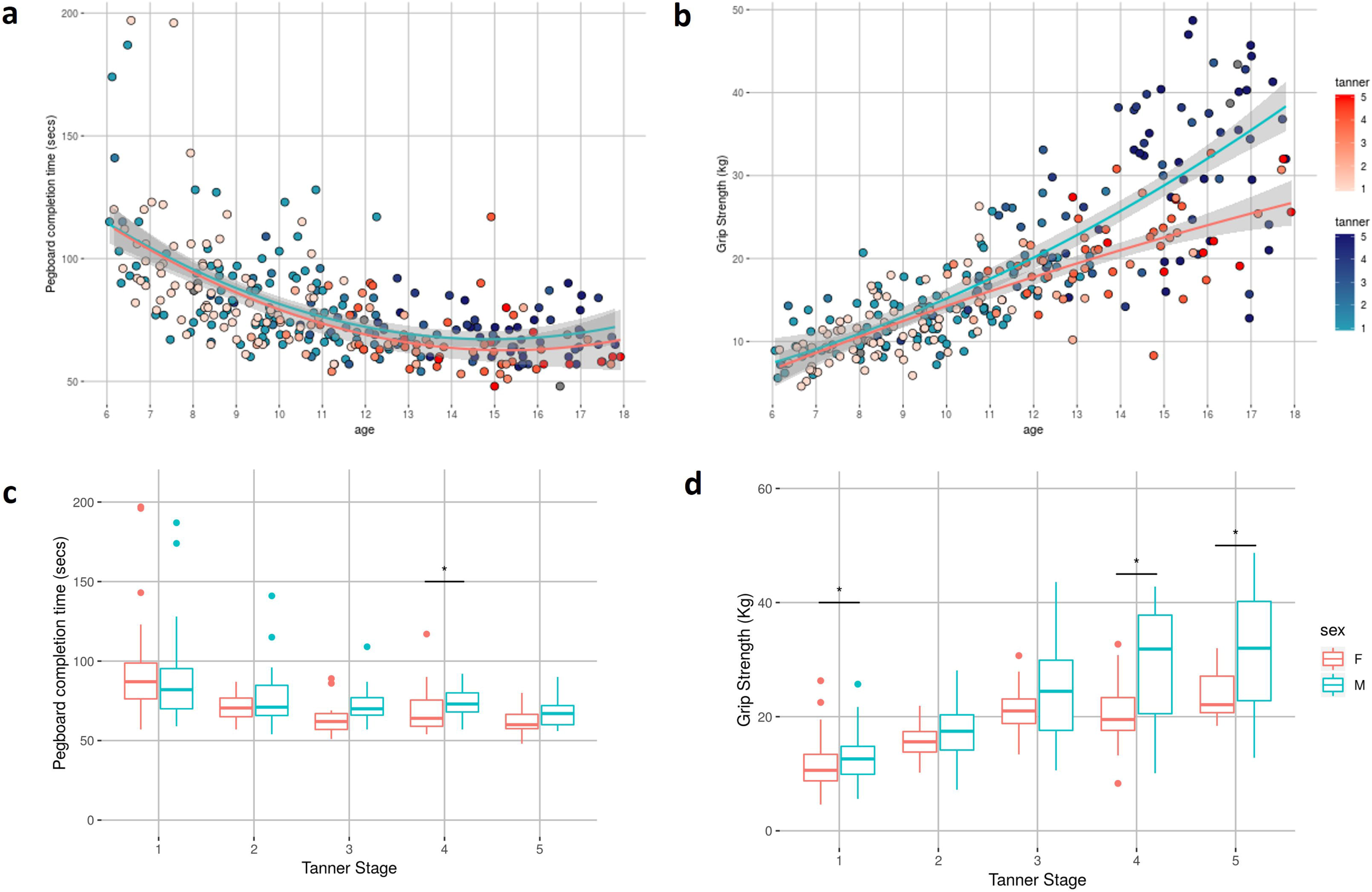
Physical measures distribution with age. (a) Time (in seconds) to completion of the Purdue Pegboard task and (b) kilogram-force Grip Strength with age are represented for all participants (age 6-17) with females coded as red and males coded as blue. Color shading degree indicates Tanner stage. Box plots by Tanner stage for (c) time to completion of Purdue Pegboard task and (d) kilogram-force Grip Strength are represented for males and females with * indicating statistical separation between males and females at that Tanner stage (p<0.05)

To further facilitate the evaluation of phenotypic data, we plotted correlations among a broad sampling of measures (see Figure 7). Statistical relationships observed after false discovery rate-based correction for multiple comparisons revealed a wealth of associations that are in general alignment with the broader psychiatric literature. For example: 1) age correlated with increased physical strength ^72^, body mass ^68^, and attention; 2) general measures of internalizing and externalizing symptoms exhibited high correlations with one another ^73–77^; 3) academic achievement correlated with intellectual aptitude ^78^ and with socioeconomic status ^79, 80^ but was inversely correlated with most internalizing and externalizing symptom measures ^81–83^—notably, ADHD traits; 4) prosocial tendencies were higher in those with lower levels of symptoms related to ADHD traits ^84^; and 5) internalizing and externalizing symptoms correlated with deficits in social cognition.

**Figure 7.**
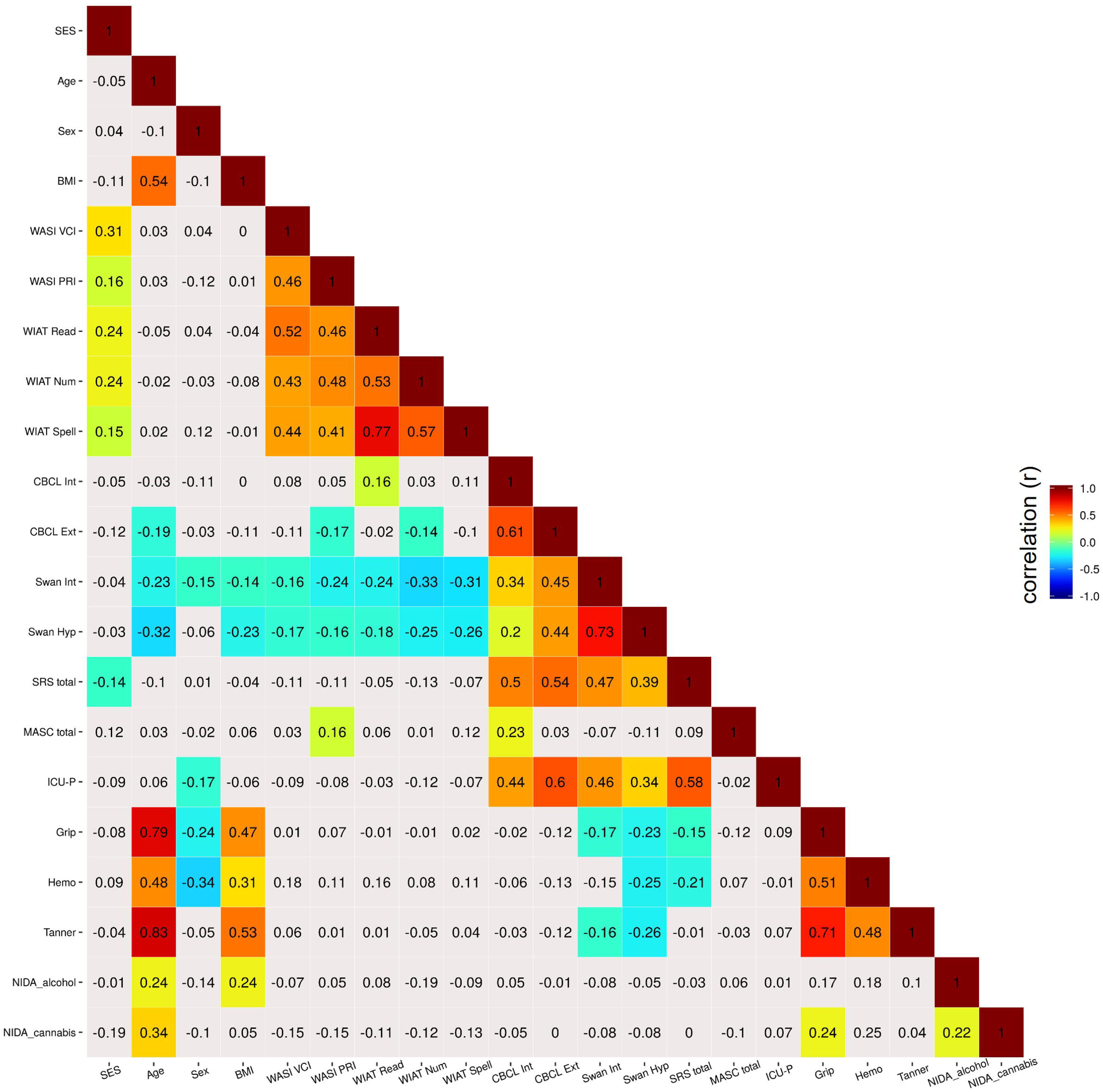
Correlation matrix of phenotypic measures. Heatmap depicting correlations between a broad sampling of behavioral, cognitive, and physical measures. Correlation values represented with color coding survived multiple comparisons correction (false discovery rate; q <0.05). BMI, Body Mass Index; WASI, Wechsler Abbreviated Scale of Intelligence-II; VCI, Verbal Comprehension Index; PRI, Perceptual Reasoning Index; WIAT, Wechsler Individual Achievement Test-II; Read, Word Reading; Num, Numerical Operations; Spell, Spelling; CBCL, Child Behavior Checklist; Int, Internalizing Subscale; Ext, Externalizing Scale; SWAN; Strengths and Weaknesses of Attention-Deficit/Hyperactivity Disorder Symptoms and Normal Behavior Scale; Hyp, Hyperactivity Subscale; SRS, Social Responsiveness Scale; MASC, Multidimensional Anxiety Scale for Children; ICU-P, Inventory of Callous-Unemotional Traits – Parent Version; Grip, Grip Strength; Hemo, Serum Hemoglobin; Tanner, Tanner Stage; NIDA, National Institute on Drug Abuse Questionnaire; Corr, Correlation.

As expected ^85^, child and parent measures assessing similar symptom domains were often poorly correlated, even for companion measures such as the Youth Self Report (YSR) and Child Behavior Checklist (CBCL) (Supplementary Figure 4). Correlation strength between the YSR and CBCL was lower than prior published data obtained in clinically referred samples ^27^. However, this difference narrowed in a post hoc subgroup analysis of participants with a baseline KSADS-PL diagnosis (Supplementary Figure 4b), arguing that YSR and CBCL are more closely correlated when psychiatric symptoms are present. Similarly, of the 121 possible intercorrelations between the YSR and CBCL syndrome scales, only two (YSR Attention Problems-CBCL Thought Problems and YSR Rule Breaking-CBCL Rule Breaking) showed statistical significance for individuals with no diagnosis at baseline (Supplementary Figure 4c). This observation is consistent with the illness orientation of clinical measures, such as YSR and CBCL and may support use of different tools for phenotyping non-clinical populations.

#### Structural MR Imaging

Measurements of T1-weighted structural images were estimated using Mindboggle, an automated morphometric analysis package that draws upon Advanced Normalization Tools (ANTS) and FreeSurfer to generate consensus outputs and an enriched set of measures for analysis ^86–90^. Brain morphometry was calculated for a total of 796 scans from 127 females and 152 males. Data from 6 females and 5 males were excluded due to insufficient quality for FreeSurfer to complete. Development curves for intracranial, gray matter, white matter, cerebral spinal fluid, and ventricular volume, mean cortical thickness, and surface area are shown in Figure 8. Data from all participants were included in these plots, independent of data quality. By using the test-retest data, reliability was calculated for each of the structural measures. Intraclass correlation coefficient (ICC) showed excellent reliability for all measures (>0.75), with cortical thickness having the lowest: Intracranial Volume = 0.99, GM Volume = 0.84, WM Volume = 0.95, CSF Volume = 0.86, Ventricular Volume = 0.99, Mean Cortical Thickness = 0.76, and mean surface area = 0.87.

**Figure 8.**
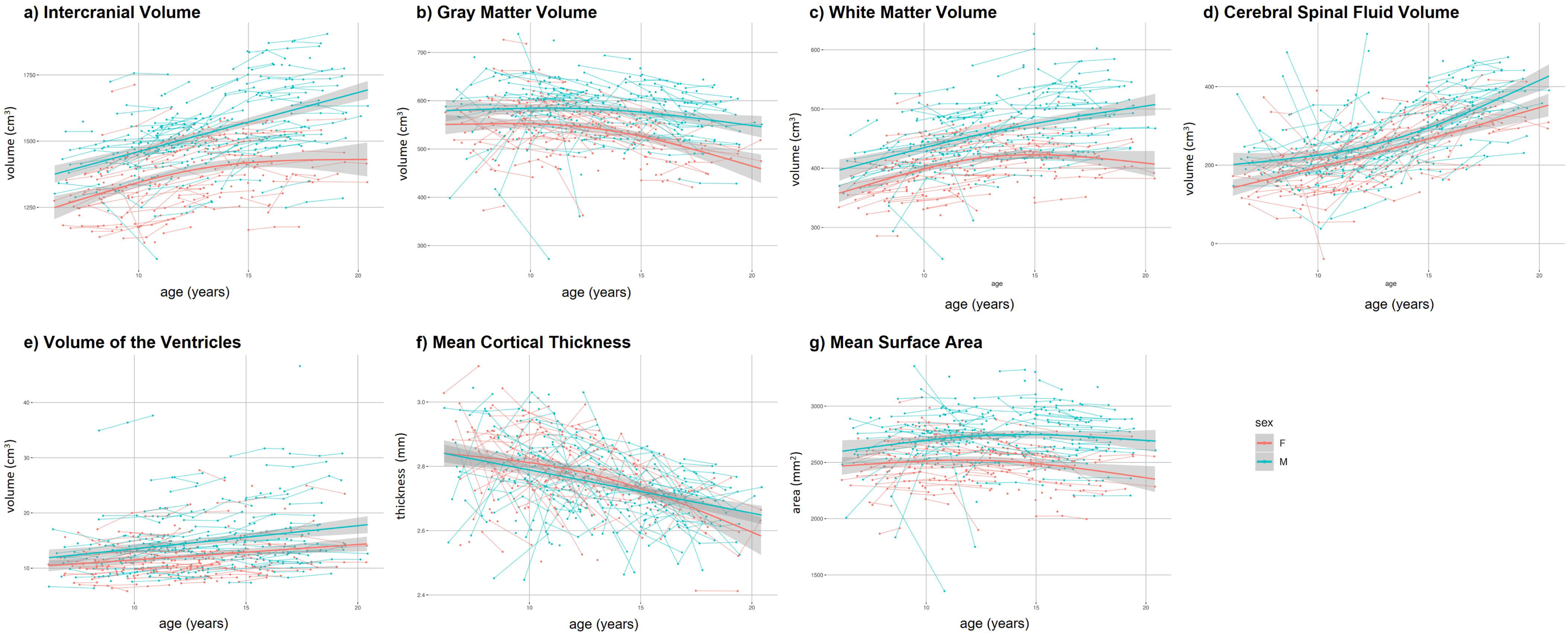
Development curves for intracranial, gray matter, white matter, cerebral spinal fluid, and ventricular volume, mean cortical thickness, and mean surface area. Data from all participants were included in these plots, independent of data quality.

To visually inspect the quality of T1-weighted images, an instance of the Braindr web application ^91^ was created. This instance contained only the images collected for the sample presented in this paper. Within Braindr, a rater has a binary choice for each image. They can either choose to pass (score = 1) or fail (score = 0) an image depending on the quality. We asked 11 raters to cast their vote based on the general quality of the image but to specifically examine if the border areas between white and gray matter are blurry or not. For each structural scan, four slices were shown to the raters (two axial and two sagittal), an approach previously used by our group ^92^. Therefore, each structural scan received 44 pass/fail votes. Slices were randomly presented to the raters. A subset of images used for training the raters is shown in Supplementary Figure 5. For each image, the average score across raters was calculated. From a total of N=585 structural scans, 455 images obtained a score greater than 0.5 (77.8%). We also compared the Braindr scores with age and Euler number, which has been shown to be a good index of structural image data quality ^92, 93^ (see Figure 9a). We found a strong age effect on the quality of structural images. For the younger participants (age <= 10 y.o.), 59.2% of the data passed quality control criteria (Braindr score >= 0.5), and for older participants (age >= 16 y.o.), 94.2% of the data passed quality control. The Receiver Operating Characteristic (ROC) curve shows that the Euler number can be reliably used as a classifier to identify passed (braindr >= 0.5) or failed (braindr < 0.5) images, with 0.91 specificity and 0.91 sensitivity. For our data, the Euler number with optimal sensitivity and specificity that identifies passable quality structural data was >-158. Of note, this Euler number might be specific to our dataset and the generalizability of the Euler number cutoff should be evaluated in future works.

**Figure 9.**
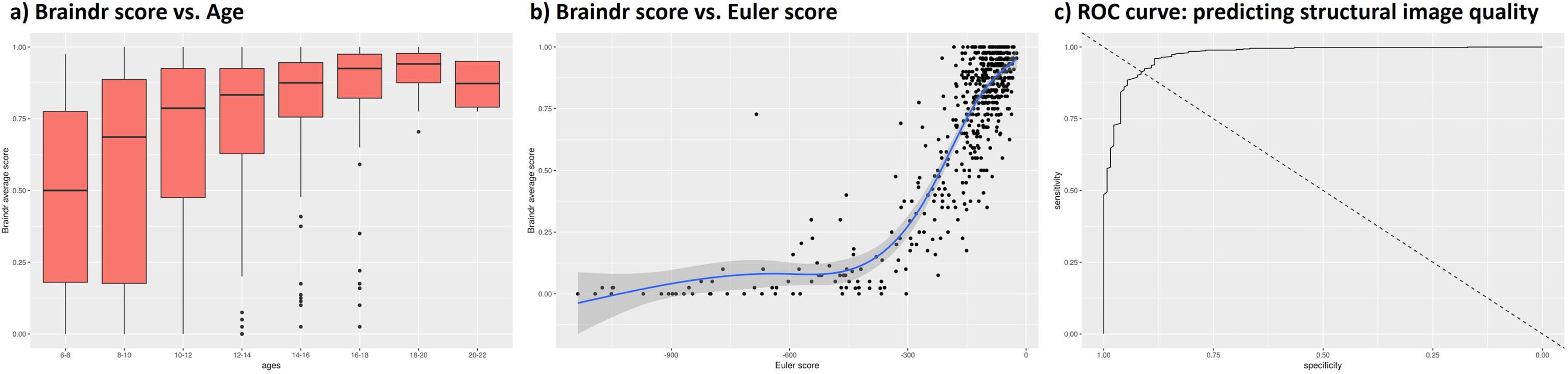
Quality control measures for structural scans. a) Average Braindr scores per age group. b) Correspondence between Braindr and Euler number. c) Receiver operating characteristic (ROC) curve showing the predictability of the Euler number to identify passed (braindr >= 0.5) or failed (braindr < 0.5) structural images.

#### Diffusion Tensor Imaging Scans

Quality control for diffusion MRI scans was calculated through the QSIprep toolbox ^94^. Specifically, the mean Framewise Displacement (FD, see Figure 10) and number of bad slices were calculated. The mean FD was 0.882 (median = 0.7181) and the mean number of bad slices was 28.57 (median = 0, range = [0, 1000]). Of note, 50.5% of the scans had zero bad slices.

**Figure 10.**
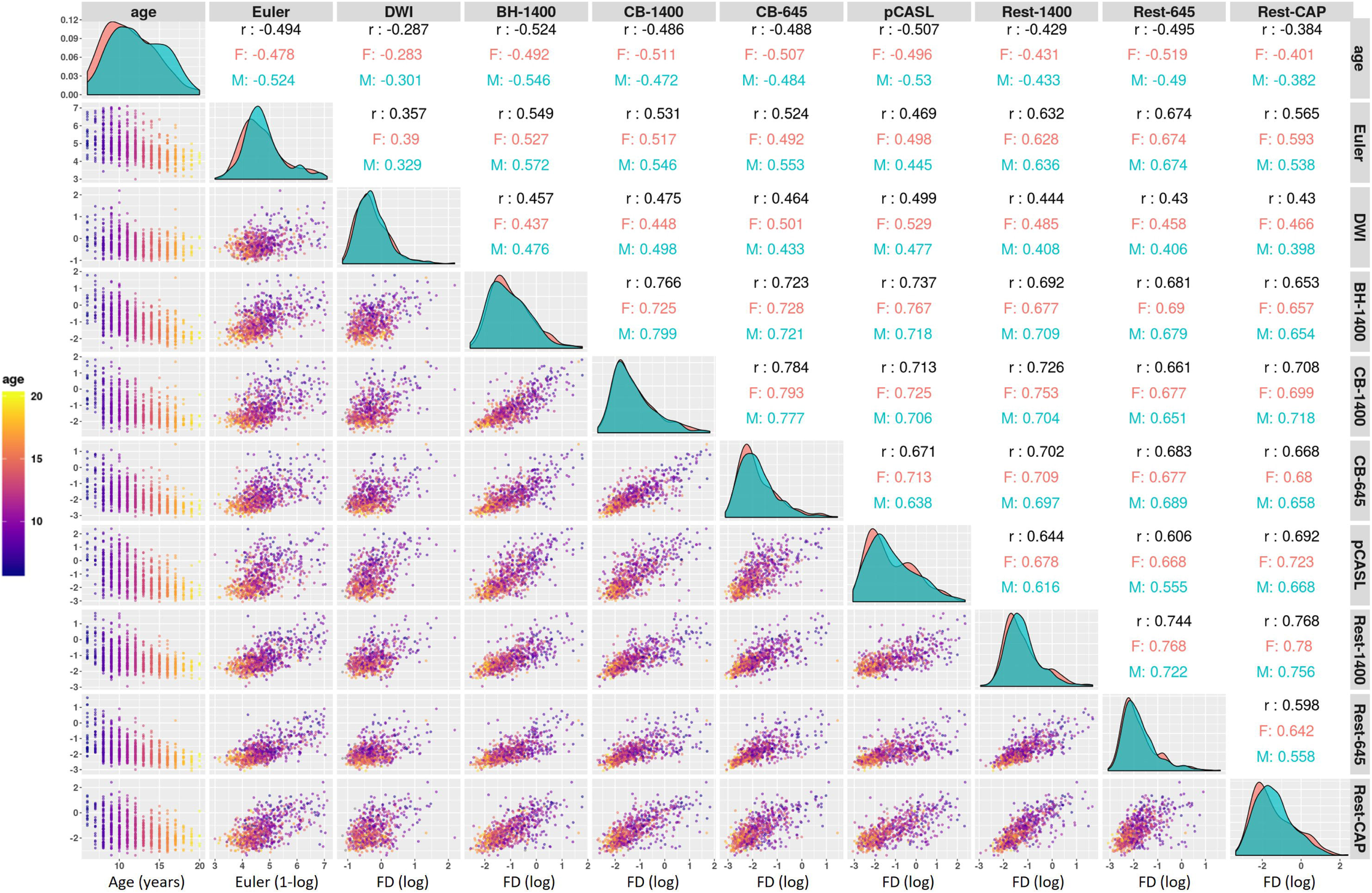
Distribution of age and quality control measures for structural scans. (Euler Number) and mean Framewise Displacement (FD) for the Diffusion Scans (DTI) and functional scans. BH-1400: Breath holding task with TR=1400ms; CB-1400: Checkerboard stimulation task with TR = 1400ms; pCASL: pseudo-Continuous Arterial Spin Labeling scan; Rest-1400: Multiband resting state scan with TR = 1400ms; Rest-645: Multiband resting state scan with TR = 645ms; Rest-CAP: Single band resting state scan with TR = 2500ms. Correlations between quality control measures are calculated across the whole sample (text in black), or within sex group (red: females; blue: males).

#### Functional Scans

To measure the relationship between age and sex with head motion, we calculated the FD ^95^ for each functional scan (Figure 10). The Euler number for the structural scans and mean FD for the diffusion scans were also included in the analysis. There is a high correlation between FD and age (r > - 0.387) for all functional scans across study participants. This high correlation holds for each sex, males and females. We also found high correlations in FD among the different functional scans (r: 0.496-0.767), as well as between the functional scans and the Euler number (r: 0.469-0.674). This high level of correlation between quality measures for different functional scans, as well as between functional and structural scans, has also been observed in other studies ^92, 96, 97^.

During the collection of fMRI, physiological data (cardiac and respiratory) were obtained during scanning. Both physiological signals are available along with the imaging data. For each participant, the mean respiration rate was calculated by identifying the peak of the Fourier spectrum of the signal. The mean respiration rate was found to be negatively correlated with age (r = -.0340), with a 18.4% reduction in rate being observed when comparing mean respiratory rates between children <=9 years old (mean = 0.347Hz) and >=17 years old (mean = 0.283 Hz), see Figure 11.

**Figure 11.**
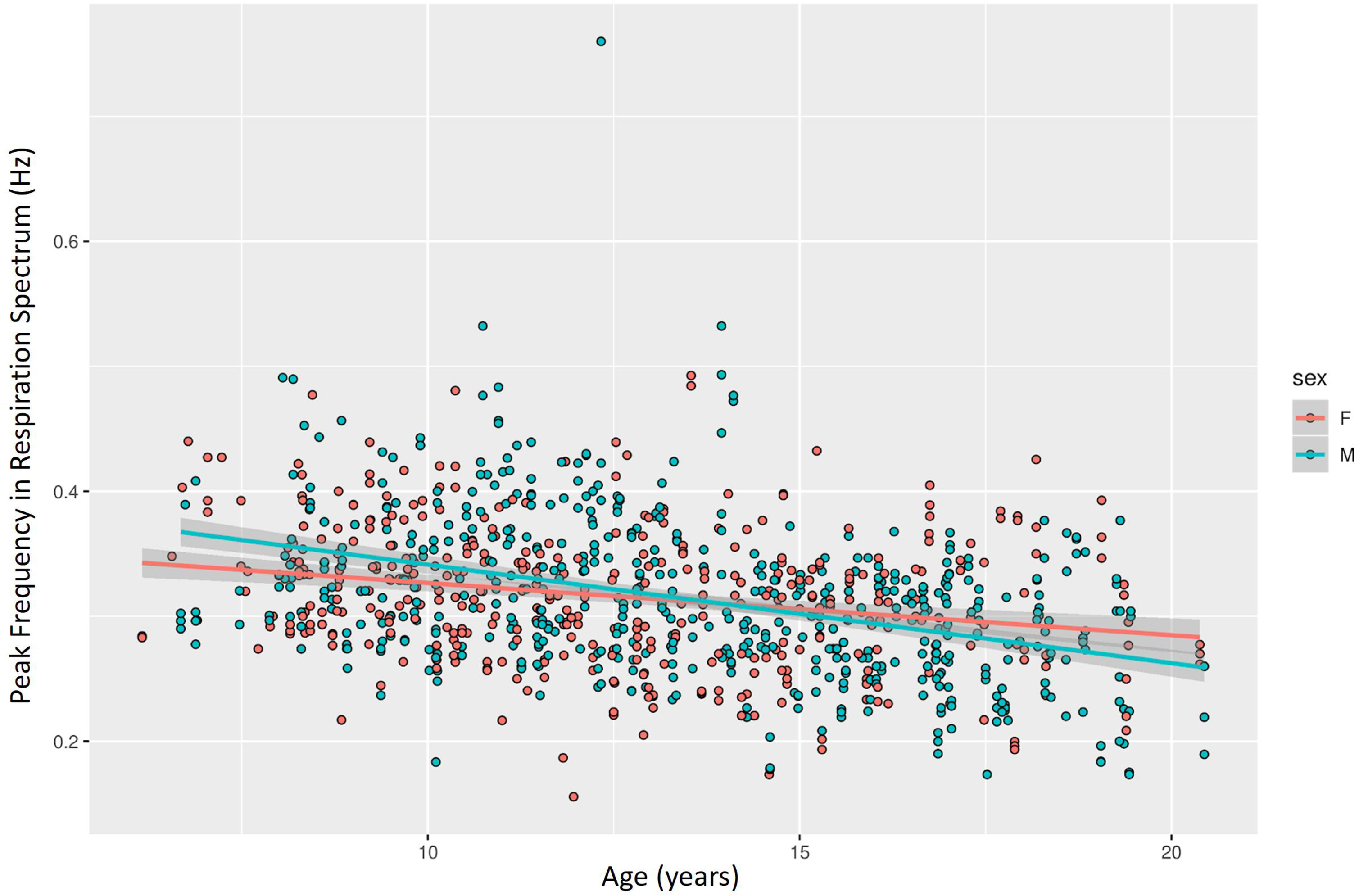
Respiration rate during fMRI scans across the sample.

## Usage Notes

*Considering Distributional Properties When Using Phenotypic Measures.* As highlighted in prior sections, several illness-focused phenotypic measures in the NKI-Rockland Sample have limited variation among individuals without symptoms. We encourage consideration of distributional characteristics of measures when formulating questions using these data. At times, it may be more sensible to limit the inclusion of participants to those with elevated levels of symptomatology (i.e., clinical, subclinical), as indicated by either general score (e.g., CBCL Total Score) or something more specific (e.g., MASC Total Score), or to treat participants without any elevations in clinical variables as a unified group. Clinical advisors are recommended members of research teams interrogating clinical phenotypic data, so they can provide guidance on relevant symptom thresholds and clinical significance of findings.

### Scaled/Standardized vs. Raw Scores

Individuals less familiar with psychiatric and cognitive assessment tools are not always aware of the transformations applied to data to account for age- and/or sex-effects. A classic example is IQ, the components of which can be present in standardized (i.e., a score of 100 represents the average IQ of the population and the standard deviation of scores is 15), scaled (e.g., average is 10; standard deviation is 3), and raw (e.g., number of items correct, number of points earned) forms. Additionally, many of the questionnaires, such as the CBCL, are transformed into a T score based on age and sex. As demonstrated under **Technical Validation**, the transformed measures for the various phenotypic tools minimize age-effects (and sex-effects, when considered); this is important because usage of the raw scores without some form of correction can result in observations being driven by age rather than other sources of individual variation.

### Cross-Initiative Harmonization

A growing number of open initiatives focused on mapping brain, cognitive and behavioral development are emerging in the community. This will create opportunities to use datasets such as the one presented here either in aggregate analysis, or as one of multiple used to establish independent replications of findings. Although promising, significant hurdles in figuring out how to put the pieces together need to be overcome before such visions can be recognized. While growing attention is being given to issues related to batch effects in imaging and differences in sampling strategies (e.g., clinically referred, community self-referred, community ascertained, community representative, enriched, etc.), differences in behavioral phenotyping protocol are often underappreciated. For example, the Adolescent Brain Cognitive Development (ABCD) ^98^; the Brazilian High Risk Cohort ^29^; the Chinese Color Nest Project (CCNP) ^99, 100^; the NIH Human Connectome Project Development (HCP-D) ^101, 102^; the Child Mind Institute Healthy Brain Network (HBN) ^103, 104^; the Pediatric Imaging, Neurocognition, and Genetics (PING) data repository ^105^; and the NKI-Rockland Sample (NKI-RS) ^16^ only share one common behavioral symptom measure—the Child Behavior Checklist ^27^—which is not included in the Philadelphia Neurodevelopmental Cohort (PNC) ^30^. In large part, this reflects differences in goals of the resources and differences in the timing of their launches (e.g., the NIH Toolbox ^106^ was not widely available when projects, such as the NKI-RS and PNC, started). Moving forward, greater focus on harmonization of samples using common data elements, and the development of methods for mapping similar though distinct tools used in different initiative samples to one another, will be crucial to facilitate cross-initiative sample aggregation and finding replication in independent datasets.

### Organization of Neuroimaging Data

Detailed instructions on how to access and download neuroimaging data are described on the study website. Briefly, neuroimaging data are organized following the Brain Imaging Data Structure (BIDS) specification ^107^. Within each participant’s folder, there are subfolders identifying the imaging sessions. Depending on participation, each participant folder may have a baseline (BAS), midpoint (Follow-up 1; FLU1), final (Follow-up 2; FLU2), and retest (TRT) subfolders. Since BIDS makes provisions for phenotypic data collected during scanning (physiological, event-related), these data are also included in each session folder in addition to the NifTI MRI series. DICOMs are not included.

### Handling Motion-Parameter Inflation by Respiration With Multiband Imaging

As has recently been noted by Power et al. ^108^ and Fair et al. ^109^, multiband functional imaging is directly affected by respiration. This artifact is most notable in the motion estimation parameters. Respiration-induced motion affects scans differently depending on scan parameters and has been observed in other large multisite neuroimaging studies that use multiband fMRI sequences, including the Human Connectome Project (HCP) and Derivatives (Lifespan ^101^ and Disease ^110^ studies) and the Adolescent Brain Cognitive Development (ABCD) ^98^ studies. None of these studies has evaluated the impact of sequences with different sampling rates on respiration-induced head motion. The resting state scans of this study with a TR = 1400ms have a much higher average framewise displacement (FD) than scans with a TR = 645ms and TR = 2500ms, with median FD_1400_ = 0.258, FD_645_ = 0.144 and FD_2500_ = 0.209, respectively (see Figure 12). The motion artifact caused by respiration is clearly noticed in the power spectrum plots of the motion parameters (Supplementary Figures 6-8), in which there is an increase in power in the 0.25-0.40 Hz range in the phase encoding direction (Anterior-Posterior). Due to aliasing, this artifact is not clearly noticed in the single-band fMRI data. To reduce this artificial respiration induced head motion, we implemented the filter approach on the motion parameters proposed by Fair et. al. A notch filter with a center frequency at 0.36 and a width of 0.07Hz was applied. However, this filter is not applicable to functional data with TR=1400 since the filter design includes the nyquist frequency of that sampling rate. Therefore, we applied a lowpass filter with a similar cutoff frequency, 0.31Hz. For the single band data (TR=2500ms), this filter is also not applicable (Nyquist frequency = 0.2Hz). We then attempted the technique suggested by Gratton et al. ^111^ and applied a low-pass filter with a cutoff frequency at 0.1Hz. Motion parameters (FD) were recalculated after applying each of the filters. By applying the 0.1Hz filter on the motion parameters, FD results are more similar across sequences. However, this method can potentially be overly aggressive by moving frames with real high motion below a specified threshold, and as such we currently recommend against its usage. Further work is still needed to compare motion estimates across different fMRI sequence parameters and evaluate preprocessing techniques to reduce these effects and their consequences in their final data analysis. These different preprocessing techniques include but are not limited to using external physiological signal correction methods (e.g., RETROICOR ^112^ and Respiratory Volume per unit Time [RVT] ^113^), COMPonent based noise CORrection (CompCor) ^114^, global signal regression ^115^, FSL FIX ^116^, among others.

**Figure 12.**
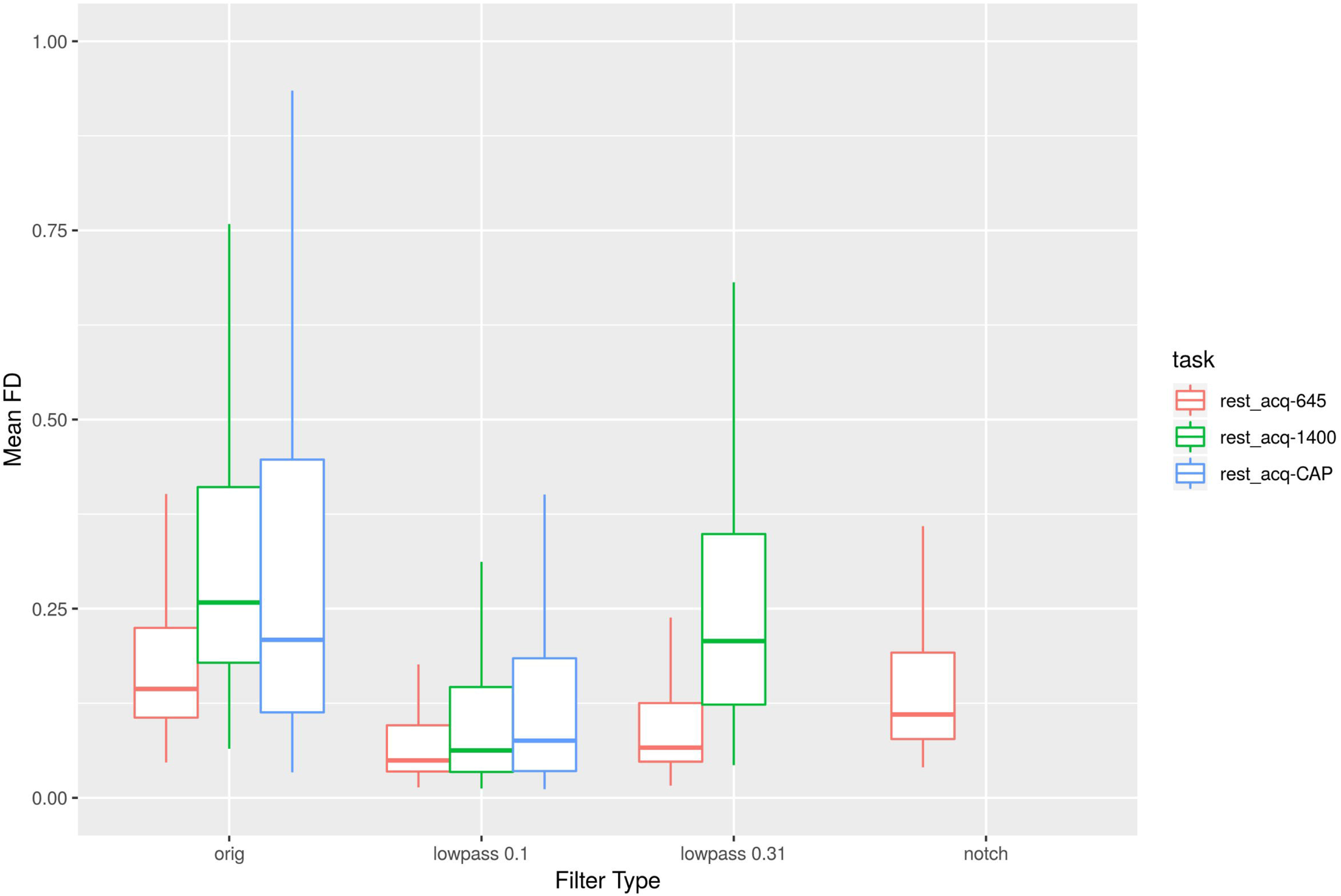
Median framewise displacement across resting state scans (TR=645/1400/2500ms) and different filtering strategies (original results, lowpass at 0.1Hz, lowpass at 0.31Hz, and notch filter with a central frequency at 0.36Hz and a width of 0.07Hz).

### Preprocessed and Derived Neuroimaging Data

In the current data release only raw neuroimaging data is being provided. Our group and others are working on discovering the optimal preprocessing strategy, focused on addressing motion induced respiration artifacts on the fMRI data mentioned above. Another problem with providing preprocessed data is that there is no one size fits all solution. For example, for some analyses, it might be optimal to perform global signal regression (e.g., reduction in distance-dependent correlation ^117^), but for others, the contrary is true (e.g., dynamic connectivity analysis ^115, 118, 119^. There is also the issue of difference in results due to preprocessing toolboxes that can have a large effect on the final analysis ^120^. Addressing all these issues/variables are beyond the scope of this data release paper. For future data releases, our group will be providing all the NKI-RS (not just of this substudy), including different preprocessing strategies, derivatives (for structural, diffusion, and functional MRI), and quality control metrics. We also encourage users to “push back” processed and derived data. Users can reach out to our group for instructions on how to upload these data to the International Neuroimaging Data Initiative (INDI) ^121^ for sharing via its AWS bucket.

## Data Availability

A more detailed overview for data access can be found at the following link: http://dx.doi.org/10.15387/fcp_indi.retro.NKIRockland. Neuroimaging data releases (http://fcon_1000.projects.nitrc.org/indi/enhanced/neurodata.html) are accessible through Amazon Web Services or NITRC. Full phenotypic data releases (http://fcon_1000.projects.nitrc.org/indi/enhanced/phenotypicdata.html) require a data use agreement.

## Acknowledgments

We thank additional team members who supported data acquisition and management: Alexis Akeyson, Ayesha Anwar, Julia Beatini, Brian Bengyak, Sinead Burrows, Jerlyne Calixte, Brian Carbone, Stephanie Carelli, Steven Carter, Tiffany Chang, Maya Charan, Danny Chon, CaraSue Doriguzzi, Jesenya DeLeon, Cornel Duhaney, Lauren Futterman, Chelsea Gessner, Alyssa Giannone, Gwen Geisler, Jamie Glass, Natacha Gordon, Courtney Gray, Caitlin Hinz, Erica Ho, Steven Homan, Olive Hwang, Christy Joseph, Stephanie Kamiel, Michelle Kaplan, Jessica Kastin, Alexis Lieval, George Lopez, Amalia McDonald, Randy Moran, Casie Morgan, Laura Panek, John Pellman, Anna Rachlin, Hayley Reed, Paula Roa, Shruti Ray, Allison Rolfe, Margaret Ryan, Sheela Sajan, Christine Santiago, Eszter Schoell, Richard Sinnig, Melissa Sital, Elise Taverna, Betty Varghese, Lauren Walden, Abigail Waters, Steven Zavitz. We are grateful to Kathleen Cuneo PhD for her clinical review and participant feedback. We thank collaborators at the TReNDS Center for insights and support in electronic data capture and distribution: Vincent D Calhoun, William Courtney, Margaret King and Dylan Wood. Additionally, we thank Alan Evans, Samir Das, and the LORIS team for the open availability of their software, as well as assistance through their support forum. We thank numerous expert consultants who contributed to the core NKI-RS protocol development: Bharat Biswal, Barbara Coffey, Christine L. Cox, Adriana Di Martino, Matthew J. Hoptman, Daniel C. Javitt, A. M. Claire Kelly, Harold S. Koplewicz, Kate Brody Nooner, Eva Petkova, Nunzio Pomara, Philip T. Reiss and John J. Sidtis. We would like to thank the AWS Open Data Sponsorship Program for storage support. We thank the numerous community partners, research participants, and families that contributed to the NKI-RS. The Longitudinal Discovery of Brain Development Trajectories was principally supported by NIH U01MH099059 (PI Milham). A subset of participants had baseline characterizations from the core enhanced NKI-RS protocol NIMH BRAINS R01MH094639-01 (PI Milham). Additional support was provided by NIH R01MH101555 (PI Craddock), NIH R01AG047596 (MPIs Milham and Colcombe), and the Child Mind Institute (1FDN2012-1). Funding for key personnel also provided in part by the New York State Office of Mental Health and Research Foundation for Mental Hygiene. Nancy Duan, Dawn Thomsen, and the Communications Team assisted in the generation of advertising materials and strategies.

## Author contributions

Conception and Experimental Design: F.X.C., S.J.C., R.C.C., V.G., B.L.L., A.M-B., M.P.M., R.H.T. Implementation and Logistics: M.B., F.X.C., S.J.C., R.C.C., V.G., M.K., B.L.L., A.M-B, M.P.M., R.H.T., K.D.T. Data Collection: M.B., S.J.C., R.C.C., C.H., M.K., A.M-B, M.P.M., R.S., R.H.T. Data Informatics: M.B., S.J.C., A.R.F., R.C.C., M.K., R.L., A.M-B, M.P.M., R.H.T. Data Analysis: M.B., S.J.C., A.R.F., M.K., R.L., A.M-B, M.P.M., R.H.T., K.D.T, L.T. Initial Drafting of the Manuscript: S.J.C., A.R.F., A.M-B, M.P.M., R.H.T. Critical Review and Editing of the Manuscript: All authors contributed to the critical review and editing of the manuscript.

## Figures

Figure images are provided as separate files.

