## Supplementary File for "A longitudinal resource for studying connectome development and its psychiatric associations during childhood"

### Supplementary Figures

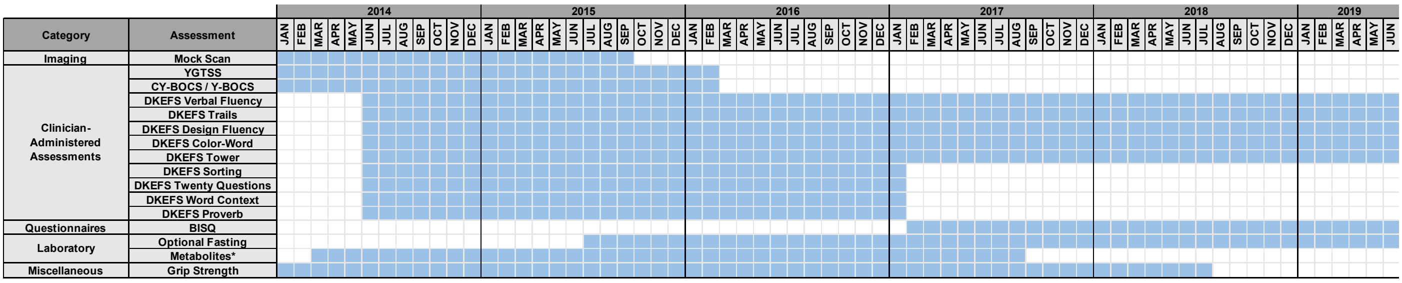

**Supplementary Figure 1. Timeline of protocol modifications.** Timeline of assessments that were added and/or removed from the study protocol. Baseline-only assessments identified in Table 2 were collected through the last participant enrollment in November 2017. BISQ = Brain Injury Screening Questionnaire; CY-BOCS = Children's Yale-Brown Obsessive Compulsive Scale; DKEFS = Delis-Kaplan Executive Function System; Y-BOCS = Yale-Brown Obsessive Compulsive Scale; YGTSS = Yale Global Tic Severity Scale. *Serum metabolites included isoleucine, kynurenine, leucine, phenylalanine, quinolinic acid, tryptophan, tyrosine, and valine. Analysis of serum metabolites was halted August 2017.

**a**

| | CLG 2-Day Baseline: Track A | | | CLG 2-Day Baseline: Track B | | |
|---|---|---|---|---|---|---|
| | Day 1 | | Day 2 | Day 1 | | Day 2 |
| 8:30 AM | Consent | | Participant Arrives/ Welcome Back | Consent | | DKEFS |
| 8:45 AM | | | | | | |
| 9:00 AM | Bloods | | MRI (MRI Time: 9:00 am) | Bloods | | |
| 9:15 AM | | | | | | |
| 9:30 AM | Breakfast | | | Breakfast | | |
| 9:45 AM | PENN CNP | KSADS | | Digit Span/WASI/WIAT | Parent Measures | |
| 10:00 AM | | | | | | |
| 10:15 AM | | | | | | |
| 10:30 AM | | | | | | |
| 10:45 AM | | | MRI-Q (age 13+), MRN | | | CYBOCS/YGTSS |
| 11:00 AM | KSADS | | | 6-Minute Bike Test | | MRN |
| 11:15 AM | Mock Scan (11:00 am) | | | Mock Scan (11:00am) | | |
| 11:30 AM | | | CYBOCS, YGTSS | | | |
| 11:45 AM | | | ANT | | | ANT |
| 12:00 PM | BIRD | | | Family History | | |
| 12:15 PM | Family History | | Dot Probe | Lunch | | Dot Probe |
| 12:30 PM | Lunch | | Lunch | | | Lunch |
| 12:45 PM | | | | BIRD | | |
| 1:00 PM | Digit Span/WASI/WIAT | Parent Measures, Vineland | DKEFS | PENN CNP | KSADS | MRI (MRI Time: 1:00 pm) |
| 1:15 PM | | | | | | |
| 1:30 PM | | | | | | |
| 1:45 PM | | | | | | |
| 2:00 PM | | | | | | |
| 2:15 PM | 6-Minute Bike Test | | | KSADS | | |
| 2:30 PM | MRN | | | MRN | Parent Measures, Vineland | MRI-Q and Satisfaction Q |
| 2:45 PM | | | | | | |
| 3:00 PM | | | Satisfaction Q | | | |
| 3:15 PM | | | Debrief | | | Debrief |

**b**

| | CLG Baseline (9:00AM MRI TIME) | | CLG Baseline (10:30AM MRI TIME) | | CLG Baseline (12:30pm MRI TIME) | |
|---|---|---|---|---|---|---|
| 8:30 AM | Re-Consent & Bloods | | Re-Consent & Bloods | | Re-Consent & Bloods | |
| 8:45 AM | | | Neuropsych | | Neuropsych | Parent Measures, Vineland, and Family Hx |
| 9:00 AM | MRI (MRI TIME: 9:00AM) | Parent Measures, Vineland, and Family Hx | | KSADS | | |
| 9:15 AM | | | | | | |
| 9:30 AM | | | | | | |
| 9:45 AM | | | MRN | | MRN | |
| 10:00 AM | | | | | | |
| 10:15 AM | | | KSADS | | | |
| 10:30 AM | MRI-Q (ages 13+) | KSADS | MRI (MRI TIME: 10:30AM) | | ANT | |
| 10:45 AM | Penn CNP | | | | | |
| 11:00 AM | | | | | Penn CNP | |
| 11:15 AM | | | | | | |
| 11:30 AM | | | | | | |
| 11:45 AM | KSADS | | | | | |
| 12:00 PM | Lunch | | MRI-Q (ages 13+) | | Lunch | |
| 12:15 PM | | | Lunch | | | |
| 12:30 PM | Neuropsych | | | | MRI (MRI TIME: 12:30PM) | |
| 12:45 PM | | | ANT | Parent Measures, Vineland, and Family Hx | | KSADS |
| 1:00 PM | | | | | | |
| 1:15 PM | | | Penn CNP | | | |
| 1:30 PM | 6-Minute Bike Test | | | | | |
| 1:45 PM | MRN | | | | | |
| 2:00 PM | | | | | MRI-Q (ages 13+) | |
| 2:15 PM | ANT | | 6-Minute Bike Test | | KSADS | |
| 2:30 PM | | | BIRD and Dot Probe | | 6-Minute Bike Test | |
| 2:45 PM | BIRD and Dot Probe | | | | BIRD and Dot Probe | |
| 3:00 PM | | | Satisfaction Q child | Satisfaction Q parent | | |
| 3:15 PM | Satisfaction Q child | Satisfaction Q parent | | | Satisfaction Q child | Satisfaction Q parent |

**c**

| Time | CLG Follow-Up (9:00AM MRI) | | CLG Follow-Up (10:30AM MRI) | | CLG Follow-Up (12:30AM MRI) | |
|---|---|---|---|---|---|---|
| 8:30 AM | Re-Consent | | Re-Consent | | Re-Consent | |
| 8:45 AM | | | Bloods | | Bloods | |
| 9:00 AM | | | | | | |
| 9:15 AM | MRI 9:00AM) | | MRN | | Digit Span/ DKEFS/ | |
| 9:30 AM | | | | KSADS | RAVLT | Parent Measures |
| 9:45 AM | | | ANT | | | |
| 10:00 AM | | | | | | |
| 10:15 AM | MRI-Q (13+) | | KSADS | | | |
| 10:30 AM | | | | | Penn CNP | |
| 10:45 AM | | | | | | |
| 11:00 AM | Penn CNP | KSADS | MRI (MRI TIME: | | | |
| 11:15 AM | | | 10:30AM) | | MRN | KSADS |
| 11:30 AM | KSADS | | | | | |
| 11:45 AM | Bloods | | | | KSADS | |
| 12:00 PM | Lunch | | MRI-Q, | | Lunch | |
| 12:15 PM | | | Lunch | | | |
| 12:30 PM | | | | | | |
| 12:45 PM | Digit Span/ DKEFS/ | | | | | |
| 1:00 PM | RAVLT | Parent Measures | Digit Span/ DKEFS/ | | MRI (MRI TIME: | |
| 1:15 PM | | | RAVLT | Parent Measures | 12:30pm) | |
| 1:30 PM | 6-Minute Bike Test | | | | | |
| 1:45 PM | ANT | | | | | |
| 2:00 PM | | | Penn CNP | | MRI-Q, | |
| 2:15 PM | | | | | ANT | |
| 2:30 PM | MRN | | | | | |
| 2:45 PM | | | BIRD and Dot Probe | | BIRD and Dot Probe | |
| 3:00 PM | BIRD and Dot Probe | | | | | |
| 3:15 PM | | | 6-Minute Bike Test | | 6-Minute Bike Test | |

**Supplementary Figure 2. Track options for ordering of assessments.** Track options include: (a) baseline two-day characterization, (b) baseline one-day characterization, and (c) follow-up visit characterization.

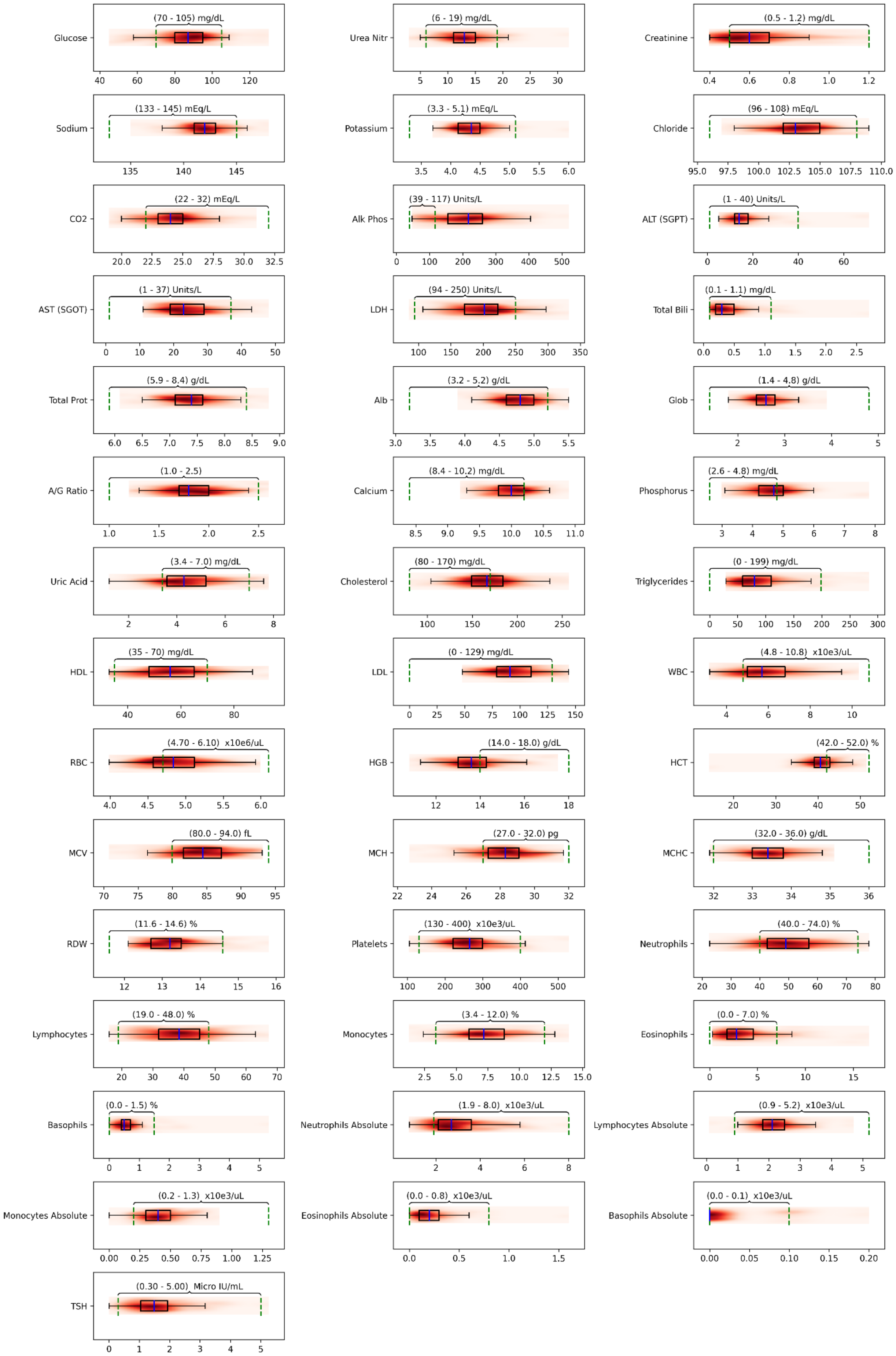

**Supplementary Figure 3. Clinical laboratory test results.** Box plots and density maps (deeper reds indicate more participants) are displayed for the study cohort. Laboratory reference ranges are adult-defined and displayed numerically with a horizontal bracket. Subjects were not all fasting before blood was drawn. The range of values for the Basophils Absolute for our sample is from 0.0 to 0.2, so all data are on the left side of the plot.

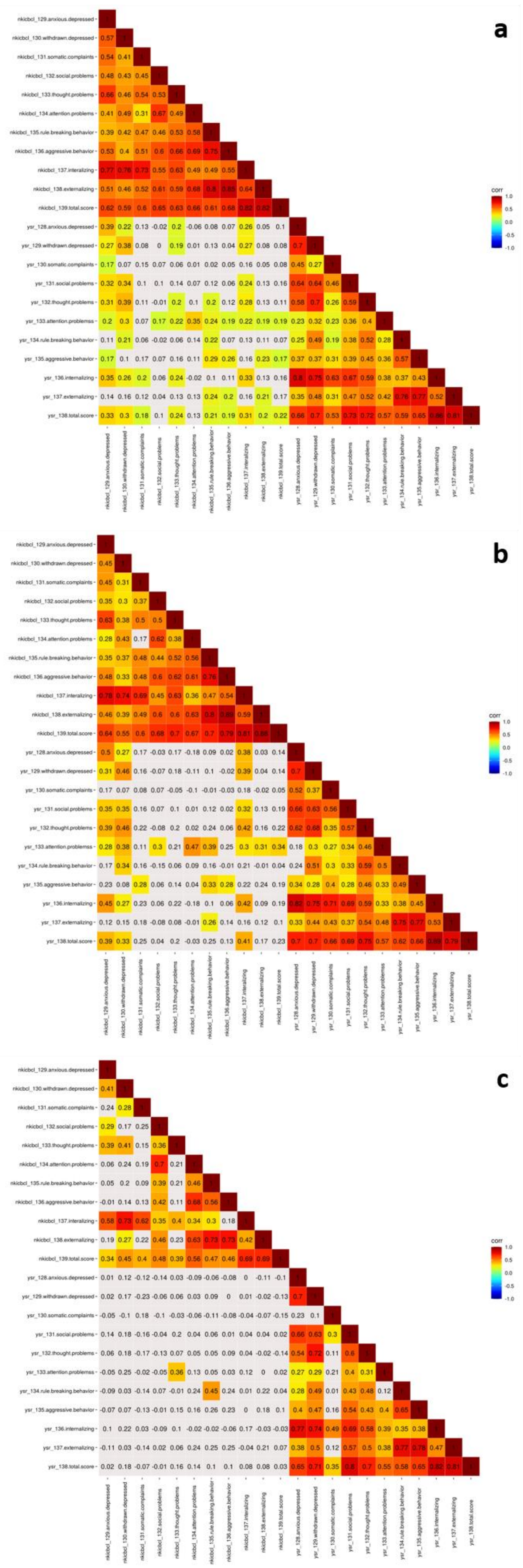

**Supplementary Figure 4. Youth Self-Report (YSR) and Child Behavior Checklist (CBCL) in behavioral phenotyping.** Heatmap depicting correlations between the Youth Self Report (YSR, a child report measure) and its companion parent report measure the Child Behavior Checklist (CBCL) in (a) all participants, (b) participants with any psychiatric diagnosis, and (c) participants with no diagnosis. Correlation values represented with color coding survived multiple comparisons correction (false discovery rate; q <0.05).

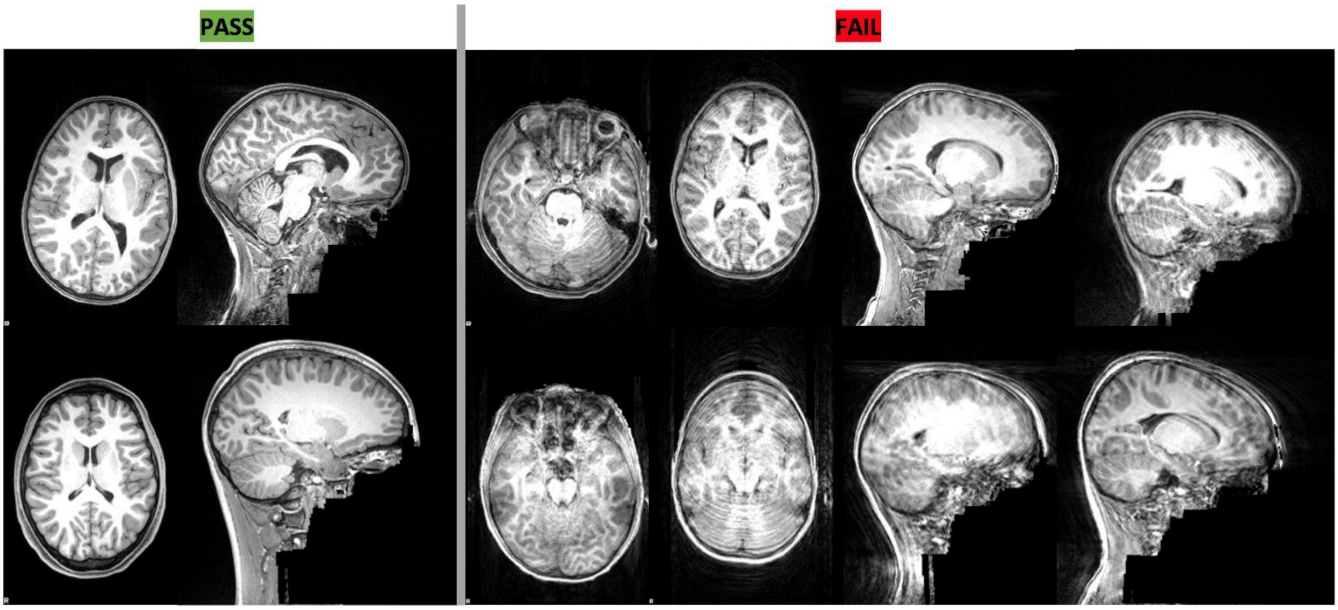

**Supplementary Figure 5. Subset of example images used for training raters to pass or fail a structural image based on quality.**

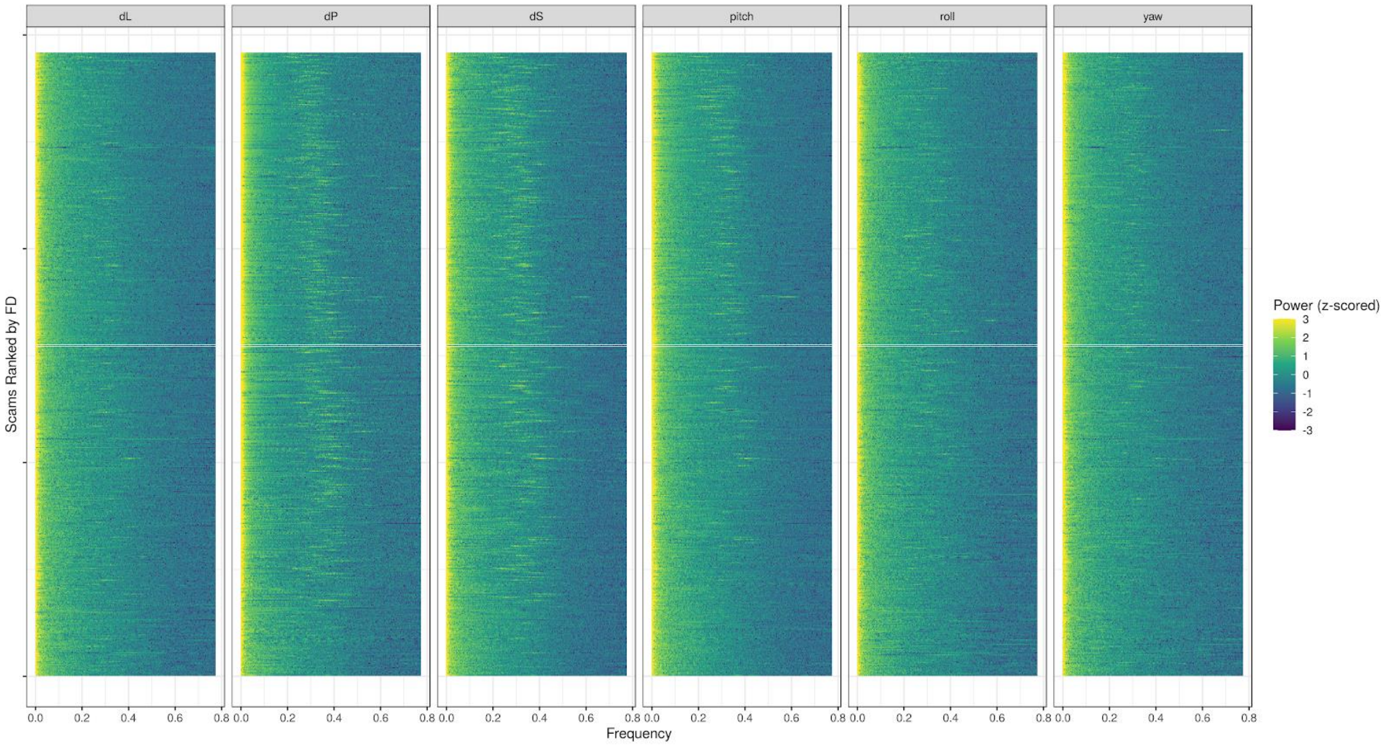

**Supplementary Figure 6. Power spectrum plots of the motion parameters of the resting state scan at TR=645ms**. Code used for plots was adapted from (Fair et al., 2020).

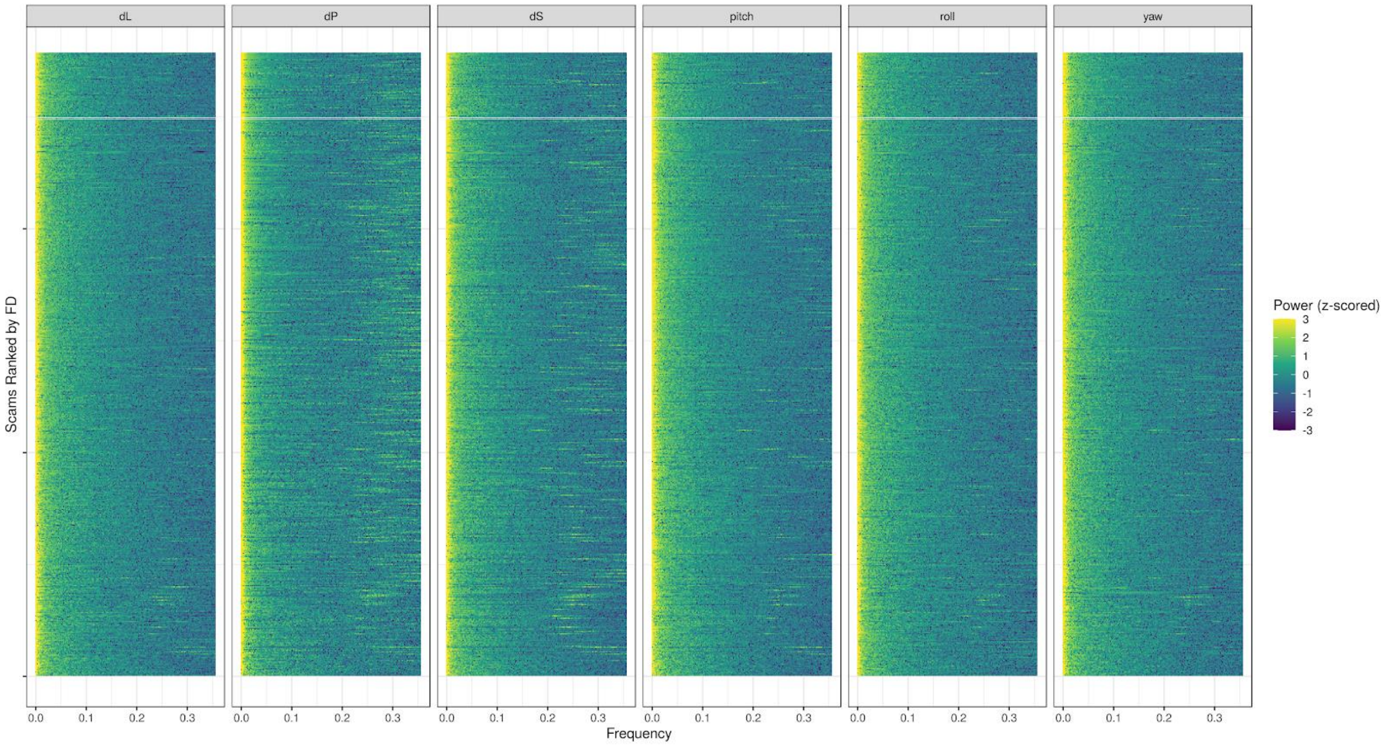

**Supplementary Figure 7. Power spectrum plots of the motion parameters of the resting state scan at TR=1400ms.** Code used for plots was adapted from (Fair et al., 2020).

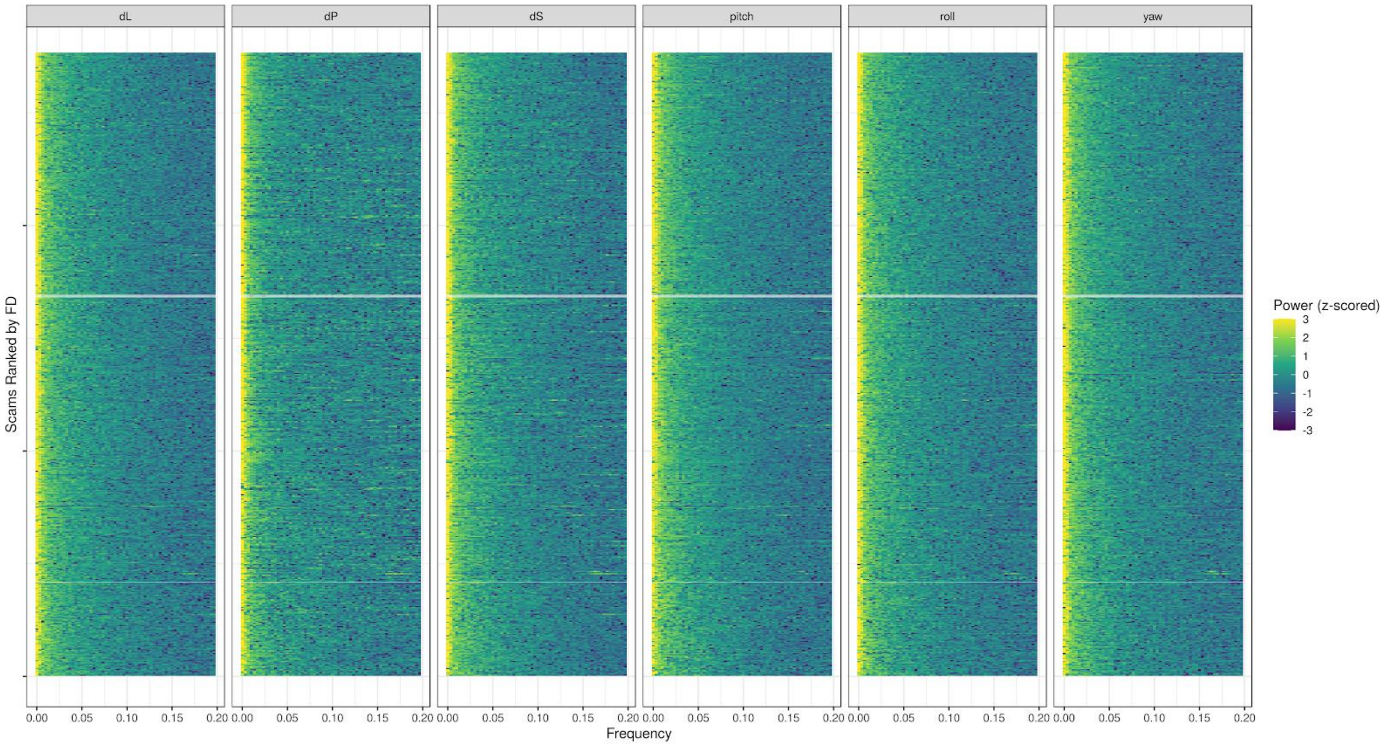

**Supplementary Figure 8. Power spectrum plots of the motion parameters of the resting state scan at TR=2500ms.** Code used for plots was adapted from (Fair et al., 2020).

#### Supplementary Tables

| Referral Source | Total | % |
|---|---|---|
| Participant Could Not Recall | 88 | 23.9% |
| Word of Mouth | 82 | 22.3% |
| Flyer | 44 | 12.0% |
| Prior NKI RS Participation | 41 | 11.1% |
| Street Fair/Community Event | 39 | 10.6% |
| NKI Neuroscience Education Day | 29 | 7.9% |
| Clinical Provider Referral | 19 | 5.2% |
| InfoUSA-based Mailings | 13 | 3.5% |
| Email/Social Media/Online | 8 | 2.2% |
| Newspaper Advertisement | 3 | 0.8% |
| Radio Advertisement | 2 | 0.5% |

**Supplementary Table 1. Primary referral pathways for enrolled participants.**

| Zip Code | Pop 2010 Census | % Pop 2010 Census | % Enrollment |
|---|---|---|---|
| **10901** | 23,465 | 7.55% | 5.52% |
| **10913** | 5,532 | 1.78% | 4.55% |
| **10920** | 8,554 | 2.75% | 2.60% |
| **10923** | 8,732 | 2.81% | 4.55% |
| **10927** | 11,910 | 3.83% | 3.25% |
| **10952** | 38,917 | 12.53% | 2.27% |
| **10954** | 23,045 | 7.42% | 8.12% |
| **10956** | 31,521 | 10.15% | 12.99% |
| **10960** | 15,093 | 4.86% | 9.42% |
| **10962** | 5,950 | 1.92% | 5.19% |
| **10964** | 1,472 | 0.47% | 0.32% |
| **10965** | 14,791 | 4.76% | 8.44% |
| **10968** | 2,353 | 0.76% | 1.30% |
| **10970** | 9,993 | 3.22% | 3.25% |
| **10974** | 3,152 | 1.01% | 0.32% |
| **10976** | 2,258 | 0.73% | 1.62% |
| **10977** | 59,048 | 19.01% | 4.55% |
| **10980** | 13,383 | 4.31% | 2.27% |
| **10983** | 5,532 | 1.78% | 7.14% |
| **10984** | 2,842 | 0.91% | 0.65% |
| **10986** | 1,974 | 0.64% | 0.97% |
| **10989** | 9,293 | 2.99% | 6.49% |
| **10993** | 4,769 | 1.54% | 1.95% |
| **10994** | 7,085 | 2.28% | 2.27% |

**Supplementary Table 2. Zip code distribution for participants residing In Rockland County.**
